# State of Emergency and Human Mobility during the COVID-19 Pandemic in Japan

**DOI:** 10.1101/2021.06.16.21259061

**Authors:** Shohei Okamoto

## Abstract

**Background:** To help control the spread of the coronavirus disease 2019 (COVID-19), the Japanese government declared a state of emergency (SoE) four times. However, these were less stringent than other nations. It has not been assessed whether soft containment policies were sufficiently effective in promoting social distancing or reducing human contact.

**Methods:** Utilising the Google mobility index to assess social distancing behaviour in all Japanese prefectures between 15 February 2020 and 21 September 2021, mobility changes were assessed by an interrupted time-series analysis after adjusting for seasonality and various prefecture-specific fixed-effects and distinguishing potential heterogeneity across multiple SoEs and time passed after the declaration.

**Results:** The mobility index for retail and recreation showed an immediate decline after the declaration of the SoE by 7.94 percent-points (95%CI: -8.77 to -7.12) and a further decline after the initial period (beta: -1.27 95%CI: -1.43 to -1.11), but gradually increased by 0.03 percent-points (95%CI: 0.02 – 0.03). This trend was similar for mobilities in other places. Among the four SoEs, the overall declines in human mobility outside the home in the third and fourth SoE were the least significant, suggesting that people were less compliant with social distancing measures during these periods.

**Conclusion:** Although less stringent government responses to the pandemic may help promote social distancing by controlling human mobilities outside the home, their effectiveness may decrease if these interventions are repeated and enforced for extended periods, distorting one’s health belief by heuristics biases. By combining these with other measures (i.e. risk-communication strategies), even mild containment and closure policies can be effective in curbing the spread of the virus.

**What is already known?:**

- Human mobility, in terms of tracing social distancing and human contact in places such as shops, restaurants, and workplaces, was reported to be a useful indicator for predicting COVID-19 outbreaks.
- Containment and closure policies, such as country lockdowns and a State of Emergency (SoE) declarations, effectively reduce human mobility.

**What are the new findings?:**

- This study first evaluated if longer and repeated SoEs were effective to reduce human mobility.
- The findings from this study suggests that although less stringent government responses to the pandemic may help promote social distancing by controlling human mobilities outside the home, their effectiveness decreases if these interventions are repeated and enforced for extended periods.

**What do the new findings imply?:**

- While less stringent government responses to the pandemic are effective in promoting social distancing by controlling human mobilities outside the home, their effectiveness may decrease if similar interventions are repeated for extended periods of time.
- However, by combining these with other measures such as risk-communication strategies, even less costly interventions such as mild containment and closure policies can be effective in curbing the spread of the COVID-19 virus.

**Research in context:** *Evidence before this study:* It has been shown that human mobility, in terms of tracing social distancing and human contact in places such as shops, restaurants, and workplaces, was reported to be a useful indicator for predicting COVID-19 outbreaks. Also, previous studies have shown that containment and closure policies, such as country lockdowns and a State of Emergency (SoE) declarations, effectively reduce human mobility. However, it is not explicitly known whether longer and repeated ‘alerts’ requesting citizens to avoid nonessential activities with risk communication strategies are equally effective.

*Added-value of this study:* This study first evaluated if longer and repeated SoEs were effective to reduce human mobility, suggesting three main findings. First, individuals engage in social distancing behaviours during the initial periods of the SoE but become less compliant as time passes. Second, when mobility changes during each SoE were distinguished, overall declines in mobilities outside the home and increases in stay-at-home time were less obvious during the succeeding SoEs. Third, under the stringent government responses to the pandemic and decline in mobilities, the consumption level—especially for activities outside the home— sharply declined, suggesting that strong public interventions may worsen the economy.

*Implications of all the available evidence:* While less stringent government responses to the pandemic are effective in promoting social distancing by controlling human mobilities outside the home, their effectiveness may decrease if similar interventions are repeated for extended periods of time. However, by combining these with other measures such as risk-communication strategies, even less costly interventions such as mild containment and closure policies can be effective in curbing the spread of the COVID-19 virus.

## Introduction

In response to the global coronavirus disease 2019 (COVID-19) health crisis, given its enormous impacts on population health, society, and economy, national governments implemented large scale public health interventions (i.e. lockdowns and state of emergency [SoE] declarations) to control the spread of the virus[1]. In addition, individuals and businesses were required to refrain from nonessential activities and to practise social distancing, leading to an economic downturn[2, 3] due to the restricted opportunities for consumption and production.

With the absence of herd immunity and/or effective treatments, minimising transmission through other means, such as wearing a mask, disinfection, ventilation, and physical distancing, are important. It has been shown that human mobility, in terms of tracing social distancing and human contact in places such as shops, restaurants, and workplaces, was reported to be a useful indicator for predicting COVID-19 outbreaks[4–8] Thus, changes in human mobility can be a tracer indicator of the general public’s response to government directives regarding the virus since this is relatively easy to capture through the use of location information (e.g. mobile phones with a Global Positioning System) as compared to other metrics (e.g. proportion of individuals wearing masks or disinfecting their hands regularly).

Previous studies have shown that containment and closure policies, such as country lockdowns and SoE declarations, effectively reduce human mobility.[9–13] However, extended restriction without complementary mitigation measures could become ineffective[14]. While tighter restriction can decrease mobility,[15, 16] it could inevitably lead to a deeper economic downturn due to the decrease in economic activities. Moreover, apart from tighter non-pharmaceutical interventions (e.g. lockdown), milder interventions such as risk communication strategies were also found to be effective in reducing the spread of infection.[15] Therefore, an ideal stringency of non-pharmaceutical interventions should be implemented, but the adopted countermeasures by each country against the pandemic highly depend on local governance and socio-economic and cultural orientations.[17, 18]

In Japan, the government responses to the pandemic are less stringent compared to other countries,[19] requesting, rather than mandating, individuals and businesses to practise social distancing and avoid nonessential activities even under a SoE. Nevertheless, human mobility was still effectively reduced in specific urban cities[9, 10]. Thus, Japan’s experiences with COVID-19 countermeasures should be helpful in supporting the mitigation of the spread of the virus by reducing human mobility with less strict interventions.

Although lockdowns and SoEs have been shown to be effective in reducing human mobility,[9–13] little is known about whether longer and repeated ‘alerts’ requesting citizens to avoid nonessential activities with risk communication strategies are equally effective. Therefore, this study aims to evaluate the association between declarations of SoEs and human mobility utilising the data covering all prefectures in Japan.

## Methods

### Data

#### (1) Human mobility

To track one of the indicators affecting COVID-19 transmissibility, a human mobility index published by Google (hereafter, mobility index) was utilised[12]. The mobility index represents the relative percentage changes from the baseline (i.e. the median value of the same day of the week between 3 January - 6 February 2020) of visits and length of stay at various places. This daily index is available for all the 47 prefectures in Japan and covers the following six categories: (a) retail and recreation; (b) supermarkets and pharmacies; (c) parks; (d) public transportation; (e) workplaces; (f) residential areas. Of these six categories, (a) retail and recreation and (c) parks could be seen as leisure activities, while (b) supermarkets and pharmacies can be considered as essential trips. Stay-at-home or work-from-home behaviours of the citizens can be seen by (d) public transport, (e) workplaces, and (f) residential. The mobility index from the baseline (between 3 January – 6 February 2020) up to 21 September 2021 was analysed in this study. The dataset includes a total of 27,495 observations from 47 prefectures and includes some missing observations for some mobility indices.

#### (2) State of Emergency

In response to the drastic virus spread, the Japanese government declared an SoE thrice in prefectures wherein the spread of infection was serious and healthcare capacities were under strain, requesting these areas to refrain from nonessential activities.

The first nationwide SoE occurred from 7 April to 25 May 2020 and was declared in seven prefectures, before expanding to all other prefectures. The second SoE was longer than the first and was declared in 11 prefectures located in urban areas from 8 January to 21 March 2021. The third SoE started on 25 April 2021 in four prefectures, and then expanded to five other prefectures. The third SoE was lifted on 20 June 2021 except for one prefecture. The fourth SoE was declared on 12 July 2021 in Tokyo, and then expanded to 17 other prefectures. The timelines of each SoE are described in Appendix Note 1.

#### (3) Policy tracker

To quantify the stringency of government responses to the pandemic, a research group from the University of Oxford published a ‘stringency index’, which comprises containment and closure policies and public information campaigns, and is reported to be associated with human mobility.[12] This index, in addition to the SoE, was used to evaluate its association with population behaviours in Japan.

As shown in Appendix Figure 1, the stringency index of Japan increased during the SoEs, and generally corresponded to the waves of infection spread.

#### (4) Household consumption

As a result of stronger government interventions to control the virus spread, the effects on the economy may be worsened due to the decrease in individual economic activity. While individual-level monthly data on economic activity is not publicly available, the Japanese government publishes monthly statistics on average household consumption, available up to July 2021, based from a nationwide sample of approximately 7,900 households[20]. Therefore, the monthly average of household consumption was used as an indicator of economic activities of the citizen. To adjust for the seasonality, percent changes from the past three-year averages of the same month were calculated. More details on the data used in this study are presented in Appendix Note 2.

### Empirical strategy

#### Detrending mobility index

The mobility index was used as the tracer indicator of the transmissibility of the coronavirus in this study. However, human mobility is largely affected by various factors, such as weather conditions, the days of the week, national holidays, and other specific events, which can differ across regions. Therefore, since simple intertemporal comparisons (e.g. pre-post and past-present comparisons) could generate biases, the mobility index could only be utilised after removing effects of weather conditions and location-specific seasonality. With this, the mobility index was detrended by obtaining the residuals (*e*_*i*,*t*_) between the actual values and the fitted values as expressed in the equation (1):

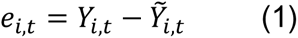

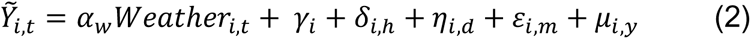

where*Y_i, t_* denotes the actual mobility index of prefecture i on the date t. 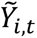 represents the linear fitted value of the mobility index predicted by daily weather conditions (i.e. mean temperature, total precipitation, sunshine duration, total snowfall, and mean wind speed), prefecture-specific fixed-effects (*γ*_*i*_), prefecture-by-national-holiday fixed-effects (*δ*_*i*,ℎ_), prefecture-by-days-of-the-week fixed-effects (*η*_*i*,*d*_), prefecture-by-month-effects (*ε*_*i*,*m*_), and prefecture-by-year fixed-effects (*μ*_*i*,*y*_). Intuitively, *e*_*i*,*t*_expresses the human mobility pattern which is not explained by weather conditions and prefecture-specific calendar effects.

#### State of Emergency and human mobility

To evaluate the association between a SoE and human mobility, several models were estimated in this study.

First, the average effects of the SoE were obtained by estimating the following equation (3):

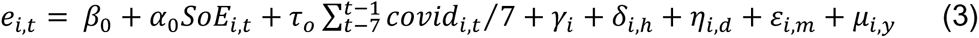

where *β*_0_ is the constant and *α*_0_ represents the parameter to be estimated for quantifying percent changes from the baseline during the SoE—a dummy variable indicating the state of emergency periods (coded 1) or not (coded 0)—of the detrended mobility index. To control potential changes in population behaviours due to the infectious conditions, the per-day infections confirmed in the past seven days for each prefecture was included in the model. The same by-prefecture fixed-effects presented in the equation (2) are also included. Moreover, the following equation (4) was estimated to assess heterogeneity in multiple SoEs:

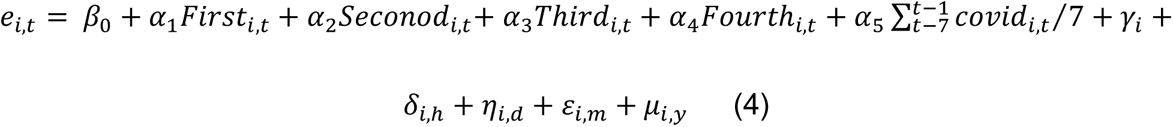

where each *α*_1_, *α*_2_, *α*_3_, and *α*_4_ represents the mobility changes in the first, second, third, and fourth SoE, respectively.

To deepen the understanding for the association between the SoE and mobility changes, an interrupted time series analysis[21] was further conducted by estimating the following equations (5):

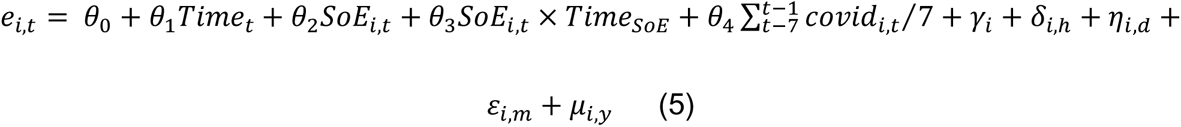

where *Time*_*t*_represents the number of days since the start of the study and *Time*_*SoE*_represents the time since the start of the SoE. Additionally, θ_1_ indicates the underlying pre-intervention trend, θ_2_ is the level of change following the SoE, and θ_3_ denotes the slope change following the SoE. By extending equation (4) in the same manner as equation (3) and (5), the mobility changes associated with each SoE was also evaluated by the time interrupted series analysis. To account for potential heteroskedasticity and autocorrelation, prefecture-level cluster robust standard errors were estimated by the panel data linear regression models with high-dimensional fixed effects[16, 22]. In addition, all estimates were weighted by the total population of each prefecture. Furthermore, potential non-linear time trends were assessed for each model, assuming a quadratic trend.

#### Government responses, mobility, and economic activity

Although the stringencies of containment and closure policies differ across regions due to differences in infectious conditions, the index was only available at the national level. Therefore, country-level analyses were conducted to assess the daily association between the stringency index and human mobility.

For consumption, given that only the monthly averages for the whole country were available, the association between the stringency index and consumption was visualised. All analyses were conducted by the Stata software version 17.0 (StataCorp LLC, College Station, USA).

## Results

### Descriptive statistics

In Table 1, descriptive statistics on key variables used in this study are presented. As the study covers all prefectures in Japan between 15 February 2020 and 21 September 2021, around 27,495 prefecture-day observations were available. Since the four seasons are distinct in Japan, the per-day mean temperature, precipitation, and sunshine duration fluctuate throughout the seasons.

**Table 1.**
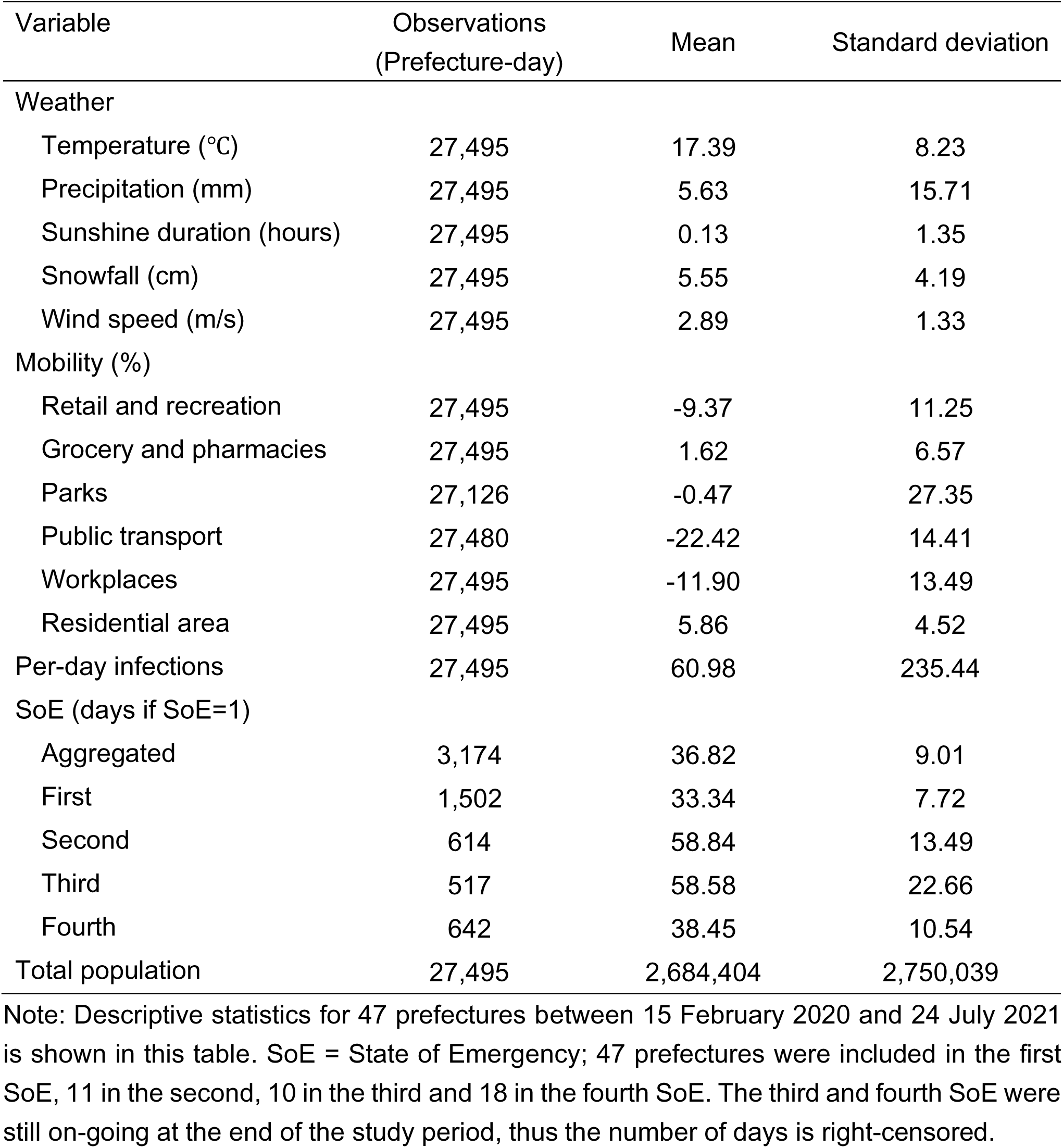
Descriptive Statistics

As for human mobility, mean percent changes from the baseline period for retail and recreation (-9.37, standard deviation [SD]: 11.25), public transport (-22.42, SD: 14.41), and workplaces (-11.90, SD: 13.49) were negative, while the one for residential was positive (5.86, SD: 4.52). Mobilities for grocery and pharmacies, and parks showed a less than or around 1% change on average.

The average duration of the first SoE was 33.34 days (SD: 7.72), taking effect in all prefectures by the end, while the other two SoEs were declared only in regions where the infectious conditions were particularly severe and healthcare capacity was under tight strain. The second SoE was the longest among the three, lasting 58.84 days on average (SD: 13.49), while the third and fourth SoE was on-going until the end of the study period, lasting 58.58 days (SD: 22.66) and 38.45 (SD: 10.54) days on average, respectively.

Figure 1 shows the descriptive presentations of detrended mobility for retail and recreation, parks, and supermarkets and pharmacies. To focus on the mobility for retail and recreation, which could be highly related to quarantine, a remarkable decline in mobility was observed during the first SoE, while it was less obvious in the succeeding SoEs. Figure 2 also presents mobilities related to self-quarantine, such as work-from-home and stay-at-home behaviours. During the first SoE, people visibly avoided using public transportation, worked from home (or stopped their businesses), and generally spent more time at home. However, the trends were found to be more unstable during the succeeding SoEs.

**Figure 1.**
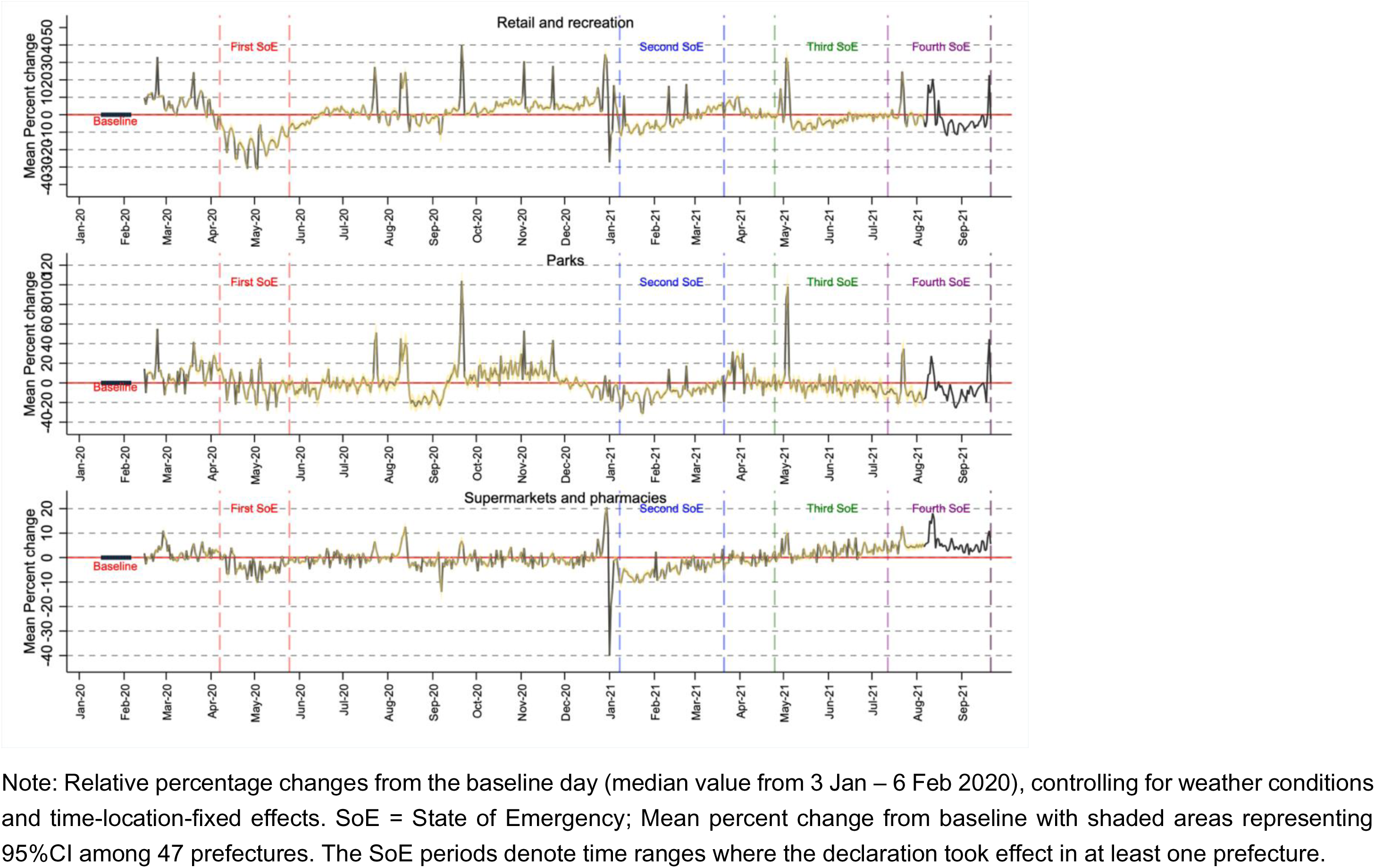
Descriptive changes in mobility: Retail and recreation, grocery and pharmacies, and parks

**Figure 2.**
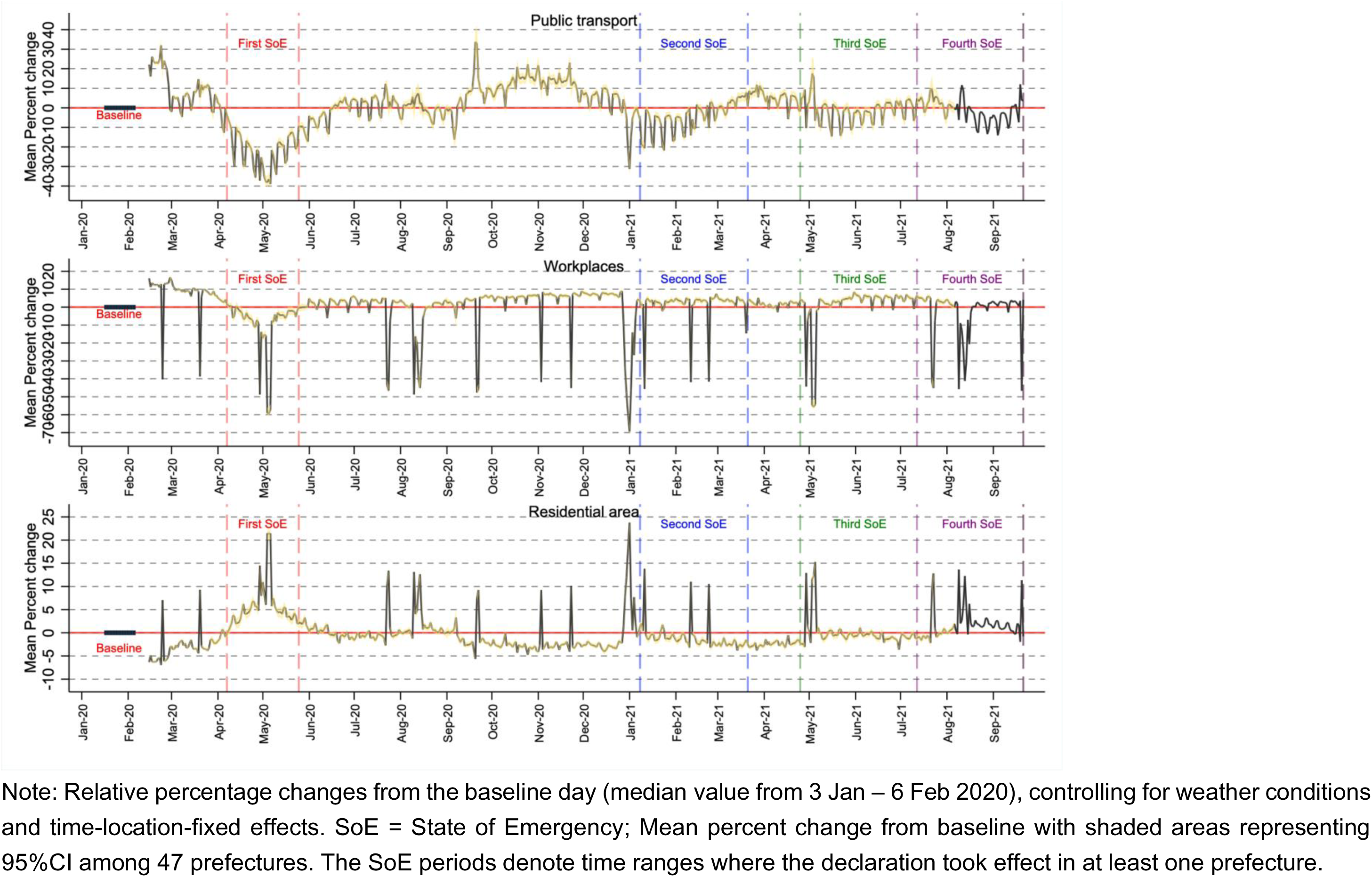
Descriptive changes in mobility: Public transport, workplaces, and residential area

### SoE and mobility

Table 2 shows the mobility changes for retail and recreation as well as residential area, associated with the SoE. Overall, visits and duration of stay at places related to retail and recreation declined during all SoE declarations, showing further drops as time passed. However, the trends (i.e. downward-convex curves) suggested that if the SoE lasted too long, it may lose its efficacy. For instance, the mobility index for retail and recreation declined by 12.88 percent-points (95% confidence interval [CI]: -13.77 to -11.99) overall during SoEs compared to non-SoE periods. The index shows an immediate drop after the declaration of the SoE by 10.10 percent-points (95%CI: -10.95 to -9.26) and a further gradual decline (beta: -0.31, 95%CI: -0.38 to -0.25). When considering the quadradic time trend, the mobility index gradually increased by 0.03 percent-points (95%CI: 0.02 – 0.03) while declines in initial periods of the SoE remained unchanged. A similar trend was observed for other outside the home mobilities for grocery and pharmacies and parks (Appendix Table 1).

**Table 2.** SoE and human mobility: Retail and recreation, grocery and pharmacies, and parks

| Model | Retail and recreation |  |  | Residential area |  |  |
| --- | --- | --- | --- | --- | --- | --- |
|  | (1) | (2) | (3) | (1) | (2) | (3) |
| Since observation |  | 0.11** | 0.09** |  | -0.02** | -0.01** |
| (day) |  | (0.10 - 0.11) | (0.07 - 0.10) |  | (-0.02 - -0.02) | (-0.02 - -0.01) |
| Since observation^2 |  |  | 0.00** |  |  | -0.00** |
| (day) |  |  | (0.00 - 0.00) |  |  | (-0.00 - -0.00) |
| SoE | -12.88** | -10.10** | -7.94** | 4.45** | 3.37** | 2.67** |
|  | (-13.77 - -11.99) | (-10.95 - -9.26) | (-8.77 - -7.12) | (4.02 - 4.88) | (2.94 - 3.80) | (2.23 - 3.12) |
| SoE days |  | -0.31** | -1.27** |  | 0.12** | 0.43** |
|  |  | (-0.38 - -0.25) | (-1.43 - -1.11) |  | (0.09 - 0.16) | (0.35 - 0.52) |
| SoE days^2 |  |  | 0.03** |  |  | -0.01** |
|  |  |  | (0.02 - 0.03) |  |  | (-0.01 - -0.01) |
| Covid cases | 0.09 | 0.02 | -0.09** | -0.01 | 0.01 | 0.05** |
| (Last week: 100) | (-0.04 - 0.21) | (-0.04 - 0.09) | (-0.13 - -0.06) | (-0.05 - 0.03) | (-0.01 - 0.03) | (0.04 - 0.06) |
| Observations | 27,495 | 27,495 | 27,495 | 27,495 | 27,495 | 27,495 |
| R-squared | 0.75 | 0.77 | 0.78 | 0.79 | 0.80 | 0.81 |
| AIC | 177,034 | 175,163 | 173,568 | 124,508 | 123,164 | 122,047 |

Note: The table presents coefficients with confidence intervals estimated from robust standard errors in parentheses. \*\* p<0.01, \* p<0.05. All models include, prefecture-level fixed-effects, prefecture-by-holiday fixed-effects, prefecture-by-weekdays fixed-effects, prefecture-by-month fixed-effects, and prefecture-by-year fixed-effects, and are weighted by population size. Constants are not presented.
SoE = State of Emergency

Additonally, people tended to use less public transport (beta: -15.31, 95%CI: -16.65 to -13.96), spend less time at workplaces (beta: -6.72, 95%CI: -7.98 to -5.47), and more time at home (beta: 4.45, 95%CI: 4.02 to 4.88) during the SoE compared to non-SoE periods (Table 2 and Appendix Table 2). The mobility index shows an immediate drop after the declaration of the SoE by 12.59 percent-points (95%CI: -14.04 to -11.13) and 4.41 percent-points (95%CI: -5.58 to -3.23) for public transport and workplaces, with a further gradual decline (beta: -0.31, 95%CI: -0.38 to -0.23; beta: -0.26, 95%CI: -0.33 to -0.20, respectively). In contrast, there was an increase by 3.37 percent-points (95%CI: 2.94 to 3.80) for residential areas, with a further gradual increase (beta: 0.12, 95%CI: 0.09 to 0.16). When considering the quadratic time trend, the mobilities for public transport and workplaces increased while that for residential area decreased as the time passed.

### Mobility during the first, second, third, and fourth SoE

Table 3 present the differences in mobility changes for retail and recreation in addition to residential area between the first, second, third, and fourth SoEs. Overall, while the mobility for retail and recreation during the first and second SoE declined, with larger magnitudes during the first SoE, the declines in mobility during the third and fourth SoEs were less significant. Although drops by 19.40 and 10.84 percent-points in the mobility for retail and recreation were observed during the first and second SoE, respectively, declines by only 3.26 and 4.60 percent-points were found in the third and fourth SoEs. Moreover, the time trends during each SoE were inconsistent. The time trends on the mobility index for retail and recreation increased during the first SoE (beta: 0.06, 95%CI: 0.02 to 0.10) and more sharply during the third SoE (beta: 0.12, 95%CI: 0.09 to 0.16), but it mildly declined during the second SoE (beta: -0.05, 95%CI: -0.09 to -0.01) and did not significantly change during the fourth SoE (beta: 0.02, 95%CI: -0.02 to 0.07). This pattern was also observed for other outside the home mobilities (Appendix Table 3 and 4).

**Table 3.**
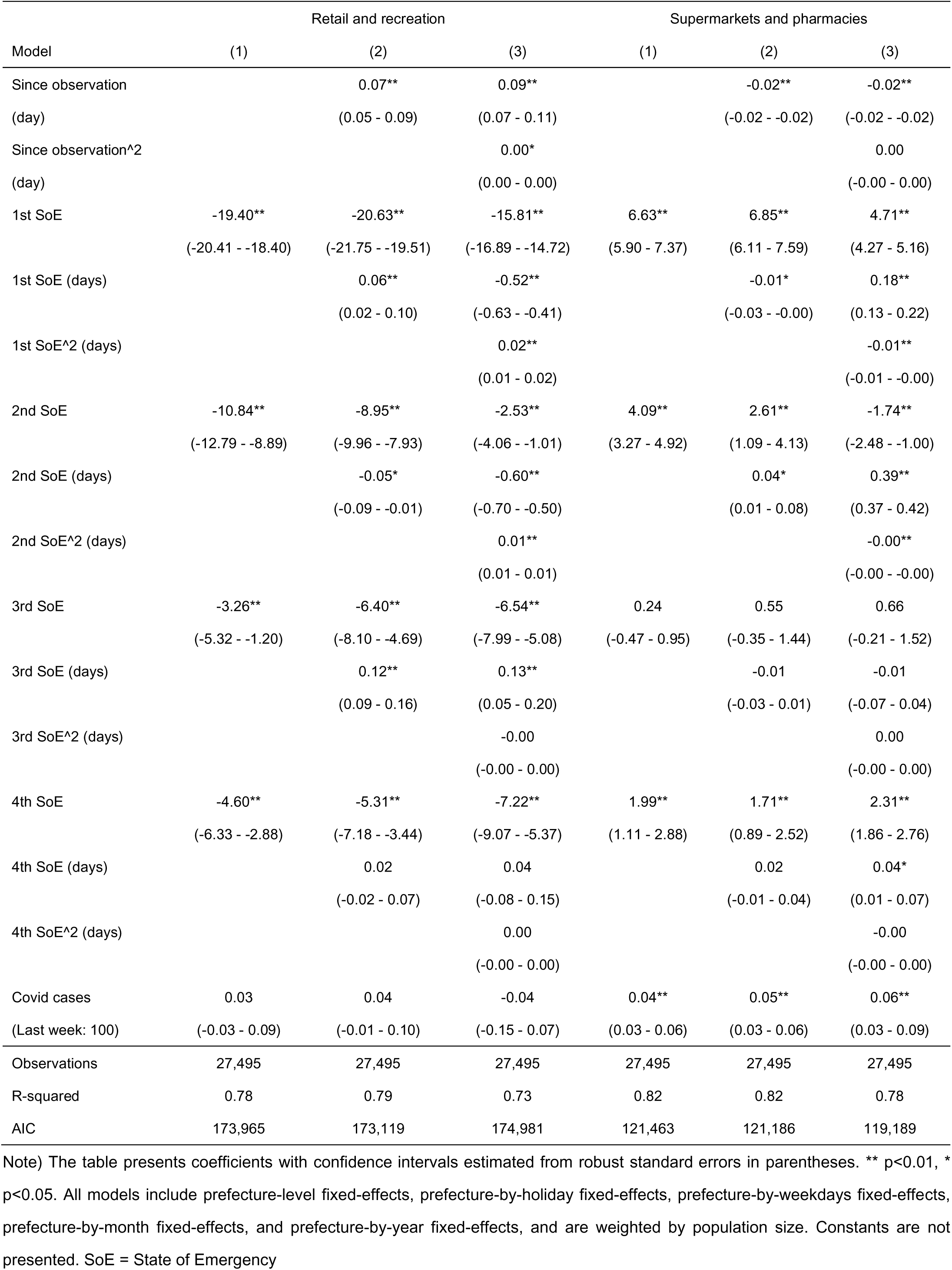
1^st^, 2^nd^, 3^rd^, and 4^th^ SoE and human mobility: Retail and recreation, grocery and pharmacies, and parks

| Model | Retail and recreation |  |  | Supermarkets and pharmacies |  |  |
| --- | --- | --- | --- | --- | --- | --- |
|  | (1) | (2) | (3) | (1) | (2) | (3) |
| Since observation |  | 0.07** | 0.09** |  | -0.02** | -0.02** |
| (day) |  | (0.05 - 0.09) | (0.07 - 0.11) |  | (-0.02 - -0.02) | (-0.02 - -0.02) |
| Since observation^2 |  |  | 0.00* |  |  | 0.00 |
| (day) |  |  | (0.00 - 0.00) |  |  | (-0.00 - 0.00) |
| 1st SoE | -19.40** | -20.63** | -15.81** | 6.63** | 6.85** | 4.71** |
|  | (-20.41 - -18.40) | (-21.75 - -19.51) | (-16.89 - -14.72) | (5.90 - 7.37) | (6.11 - 7.59) | (4.27 - 5.16) |
| 1st SoE (days) |  | 0.06** | -0.52** |  | -0.01* | 0.18** |
|  |  | (0.02 - 0.10) | (-0.63 - -0.41) |  | (-0.03 - -0.00) | (0.13 - 0.22) |
| 1st SoE^2 (days) |  |  | 0.02** |  |  | -0.01** |
|  |  |  | (0.01 - 0.02) |  |  | (-0.01 - -0.00) |
| 2nd SoE | -10.84** | -8.95** | -2.53** | 4.09** | 2.61** | -1.74** |
|  | (-12.79 - -8.89) | (-9.96 - -7.93) | (-4.06 - -1.01) | (3.27 - 4.92) | (1.09 - 4.13) | (-2.48 - -1.00) |
| 2nd SoE (days) |  | -0.05* | -0.60** |  | 0.04* | 0.39** |
|  |  | (-0.09 - -0.01) | (-0.70 - -0.50) |  | (0.01 - 0.08) | (0.37 - 0.42) |
| 2nd SoE^2 (days) |  |  | 0.01** |  |  | -0.00** |
|  |  |  | (0.01 - 0.01) |  |  | (-0.00 - -0.00) |
| 3rd SoE | -3.26** | -6.40** | -6.54** | 0.24 | 0.55 | 0.66 |
|  | (-5.32 - -1.20) | (-8.10 - -4.69) | (-7.99 - -5.08) | (-0.47 - 0.95) | (-0.35 - 1.44) | (-0.21 - 1.52) |
| 3rd SoE (days) |  | 0.12** | 0.13** |  | -0.01 | -0.01 |
|  |  | (0.09 - 0.16) | (0.05 - 0.20) |  | (-0.03 - 0.01) | (-0.07 - 0.04) |
| 3rd SoE^2 (days) |  |  | -0.00 |  |  | 0.00 |
|  |  |  | (-0.00 - 0.00) |  |  | (-0.00 - 0.00) |
| 4th SoE | -4.60** | -5.31** | -7.22** | 1.99** | 1.71** | 2.31** |
|  | (-6.33 - -2.88) | (-7.18 - -3.44) | (-9.07 - -5.37) | (1.11 - 2.88) | (0.89 - 2.52) | (1.86 - 2.76) |
| 4th SoE (days) |  | 0.02 | 0.04 |  | 0.02 | 0.04* |
|  |  | (-0.02 - 0.07) | (-0.08 - 0.15) |  | (-0.01 - 0.04) | (0.01 - 0.07) |
| 4th SoE^2 (days) |  |  | 0.00 |  |  | -0.00 |
|  |  |  | (-0.00 - 0.00) |  |  | (-0.00 - 0.00) |
| Covid cases | 0.03 | 0.04 | -0.04 | 0.04** | 0.05** | 0.06** |
| (Last week: 100) | (-0.03 - 0.09) | (-0.01 - 0.10) | (-0.15 - 0.07) | (0.03 - 0.06) | (0.03 - 0.06) | (0.03 - 0.09) |
| Observations | 27,495 | 27,495 | 27,495 | 27,495 | 27,495 | 27,495 |
| R-squared | 0.78 | 0.79 | 0.73 | 0.82 | 0.82 | 0.78 |
| AIC | 173,965 | 173,119 | 174,981 | 121,463 | 121,186 | 119,189 |

Note) The table presents coefficients with confidence intervals estimated from robust standard errors in parentheses. \*\* p<0.01, \* p<0.05. All models include prefecture-level fixed-effects, prefecture-by-holiday fixed-effects, prefecture-by-weekdays fixed-effects, prefecture-by-month fixed-effects, and prefecture-by-year fixed-effects, and are weighted by population size. Constants are not presented. SoE = State of Emergency

More visits and time spent at residential areas were not observed during the third SoE, although this was observed during the first, second, and fourth SoEs. Furthermore, the immediate increase in stay-at-home time was the largest in the first SoE (beta: 6.85, 95%CI: 6.11 to 7.59). For the second SoE, a modest initial increase in stay-at-home time by 2.61 percent-points (95%CI: 1.09 to 4.13) was followed by a gradual increase (beta: 0.04, 95%CI: 0.01 to 0.08), while no change was observed during the third SoE. During the fourth SoE, a modest increase was observed (beta: 1.71, 95%CI: 0.89 to 2.52).

### Government responses, mobility, and consumption

In addition to mobility changes associated with the SoE, the associations between mobility indices and stringency of government responses to the pandemic were assessed (Appendix Table 5 and 6). With tighter government responses, most of the mobility outside the home declined while residential mobility increased. Decomposing containment and closure policies, school closing, workplace closing, public transport closing, stay-at-home orders were associated with the declined mobility for retail and recreation and the increased residential mobility. In contrast, the associations between restrictions on gatherings and mobilities at these places were opposite.

Moreover, although the consumption data during the fourth SoEs were limitedly available, the per-month total consumption declined by about 13.2% and 7% in the first and second SoE, respectively, compared to the past-three-year average (Appendix Figure 2). Consumptions, especially for public transportation, culture and recreation, and clothing and footwear—dimensions which can be linked to activities outside the home—remarkably declined at most by about 20%, 35%, and 57% between April and May in 2020 during the first SoE, about 11%, 21%, 33% between January and March 2021 during the second SoE, and about 1%, 19%, 28% in May 2021 during the third SoE.

### Robustness checks

Further analyses were conducted to evaluate mobility changes associated with a quasi-SoE, a semi-state of emergency was implemented by the Japanese government, reflecting the infectious conditions and their effects on healthcare capacity (Appendix Tables 7-10). However, the quasi-SoE was only weekly or non-significantly associated with mobility changes to reduce human contact.

Mobility changes of the population may differ across regions depending on the infectious conditions. For instance, citizens in prefectures with higher numbers of infections may be more likely to engage in social distancing compared to those with lower numbers. Therefore, the prefectures were classified into two groups: High and Low. A prefecture was classified as ‘High’ if the cumulative number of confirmed cases in that area was above the median of all prefectures within the study period. In contrast, ‘Low’ prefectures were those with median or below median confirmed cases. In both groups, mobility patterns were similar, showing a decline in activities outside the home and an increase in stay-at-home behaviours after the declaration of an SoE (Appendix Tables 11 and 12). However, the post-SoE slopes suggest conflicting trends: a gradual decrease in mobilities for retail and recreation and an increase in residential visits and duration were observed in the ‘High’ group, while the opposite trend was found in the ‘Low’ counterpart. When considering non-linear time trends, identical results were found for both groups. Findings for mobility changes during each SoE for the ‘High’ group were almost identical with the pooled estimates for all prefectures (Appendix Tables 13 and 14).

Moreover, the trust in the government, measured by the approval rating for the cabinet, may attenuate the association between the SoE and mobility under the state where the citizens are requested to follow the guideline. However, this was not evident (Appendix Table 15). Additionally, the association between vaccination rates and mobility was analysed, showing that vaccination rates were positively associated with the mobility for retail and recreation regardless of the declaration of SoE (Appendix Table 16).

## Discussion

This study aimed to evaluate mobility changes during the SoE after distinguishing potential heterogeneity across each SoE and time passed after each declaration. To summarise, three main findings were found in this study.

First, human mobility in places outside the home effectively declined while stay-at-home time increased during SoEs. This was also confirmed by adopting the interrupted time-series analysis, suggesting that individuals engage in social distancing behaviours during the initial periods of the SoE but become less compliant as time passes.

Second, when mobility changes during each SoE were distinguished, overall declines in mobilities outside the home and increases in stay-at-home time were less obvious during the succeeding SoEs. For instance, during the third and fourth SoE, immediate declines in mobilities were smaller or not found in all places.

Third, under the stringent government responses to the pandemic and decline in mobilities, the consumption level—especially for activities outside the home—sharply declined, suggesting that strong public interventions may worsen the economy.

Under a state where people are requested to voluntarily engage in social distancing behaviours, it may be difficult for them to comply for extended and repeating periods. Following the Health Belief Model[23], individual beliefs regarding the virus (e.g. susceptibility and severity), benefit of one’s behaviours, barriers to make the desired behavioural changes, and self-efficacy to overcome given challenges affect an individual’s actual social distancing behaviour. By being subjected to a SoE multiple times, individuals could become more optimistic to the infection due to heuristics biases[24], which may lead individuals to engage in higher-risk behaviours.

Even though the mobility indices were detrended by weather conditions and various prefecture-specific fixed-effects, there can be notable differences between the first, second, third, and fourth SoE. In particular, the third SoE occurred during one of the busiest holiday seasons in Japan (i.e. Golden Week), which could be a factor in motivating individuals to leave their homes. Furthermore, the fourth SoE was declared in the Olympic Games and the summer break season. Thus, the mobility level may not be at the expected level for a pandemic and SoE even though it might have been well-suppressed during the pre-pandemic Golden Weeks and summer breaks. Additionally, the fear of new coronavirus variants with potentially high transmissibility rates and disease severity could have also affected population behaviours. Moreover, while the association between the national-level government responses to the pandemic and mobility was analysed in this study, the contents of requests by the local government were not identical for each prefecture, which may partly contribute to different mobility patterns across regions. Thus, further studies are demanded to evaluate this to obtain insights on effective responses to the pandemic.

Vaccinated individuals with greater immunity may be less willing to engage in social distancing behaviours. In fact, findings of this study suggest that vaccination rates were associated with the increase in mobility for retail and recreation. However, the government of Japan continues to request individuals, even after fully vaccinated, to be compliant with social distancing. Therefore, assisting the progress toward herd immunity against COVID-19 by increasing vaccination uptake may be more efficient, rather than relying on non-pharmaceutical interventions when a vaccine is available.

Until the majority of the population is fully vaccinated allowing for herd immunity, it is still vital to promote individual social distancing behaviours, such as wearing a mask, disinfection, and avoiding mass gatherings. Considering that stringent and widescale public health interventions could damage the economy, and the fact that repeating the same interventions could become ineffective as shown in this study, other appropriate measures need to be enforced in order to curb the virus spread. It has been shown that even less costly interventions, such as risk-communication strategies, can be considered as highly effective.[15] Therefore, by utilising behavioural insights[26], methods to deliver directives to the population should be carefully designed to maximise their effectiveness.

Despite the significant findings of this study, there are still some limitations that should be considered. First, the Google mobility index used in the study is limited since it does not count an absolute amount of human mobility. Although the mobility index was detrended by weather conditions and various prefecture-specific fixed-effects, relative changes from the baseline given narrow time range may be inappropriate, making interpretation difficult. Also, the Google mobility index does not include information on smaller units of regions (e.g. city-level), time (e.g. daytime and night), and comparisons by sex and age. Second, the generalisability of the findings should be carefully considered. As the government responses to the pandemic could reflect cultural and institutional differences[17, 18], the findings from this study may not be applicable in other countries. Third, as mentioned earlier, the contents of requests by the local governments varied. However, potential heterogeneity in effectiveness of each restriction by prefecture was not assessed in this study as it was beyond the scope of this study. Nevertheless, implications from this study should be helpful in highlighting the limitations of less stringent, repeated, and prolonged public health interventions.

In conclusion, while less stringent government responses to the pandemic are effective in promoting social distancing by controlling human mobilities outside the home, their effectiveness may decrease if similar interventions are repeated for extended periods of time. However, by combining these with other measures such as risk-communication strategies, even less costly interventions such as mild containment and closure policies can be effective in curbing the spread of the COVID-19 virus.

## Data Availability

All data are obtained from publicly available sources.

## Ethics

Ethical approval was not required as this study was based on secondary analysis of publicly available data.

## Supplementary Material

### Appendix Note 1: Time line of the state of emergency in Japan

The time line of the first SoE

- 7 April 2020: Took effect in Saitama, Chiba, Tokyo, Kanagawa, Osaka, Hyogo, and Fukuoka
- 16 April 2020: Expanded to all prefectures
- 14 May 2020: Lifted in 39 prefectures except for Hokkaido, Saitama, Chiba, Tokyo, Kanagawa, Osaka, Kyoto, and Hyogo
- 21 May 2020: Lifted in Osaka, Kyoto, and Hyogo
- 25 May 2020: Lifted in remaining 5 prefectures

The time line of the Second SoE

- 8 January 2021: Took effect in Saitama, Chiba, Tokyo, and Kanagawa
- 14 January 2021: Took effect in Tochigi, Gifu, Aichi, Kyoto, Osaka, Hyogo, and Fukuoka
- 8 February 2021: Lifted in Tochigi
- 8 March 2021: Lifted in Gifu, Aichi, Kyoto, Osaka, Hyogo, and Fukuoka
- 22 March 2021: Lifted in Saitama, Chiba, Tokyo, and Kanagawa

The time line of the Third SoE

- 25 April 2021: Took effect in Tokyo, Kyoto, Osaka, and Hyogo
- 12 May 2021: Took effect in Aichi and Fukuoka
- 16 May 2021: Took effect in Hokkaido, Okayama, and Hiroshima
- 23 May 2021: Took effect in Okinawa
- 20 June 2021: Lifted in Hokkaido, Tokyo, Aichi, Kyoto, Osaka, Hyogo, Okayama, Hiroshima, and Fukuoka
- 30 September 2021: Scheduled to be lifted in Okinawa

The time line of the Fourth SoE

- 12 July 2021: Took effect in Tokyo
- 2 August 2021: Took effect in Saitama, Chiba, Kanagawa, and Osaka
- 20 August 2021: Took effect in Ibaraki, Tochigi, Gumma, Shizuoka, Kyoto, Hyogo, and Fukuoka
- 27 August 2021: Took effect in Hokkaido, Gifu, Aichi, Mie, Shiga, and Hirosima
- 30 September 2021: Scheduled to be lifted

### Appendix Note 2: Data used in this study

#### Human mobility index

This index was obtained from Google LLC[1], which indicates relative changes from a baseline in visits and length of stay at different places. The index was created by the location history of individuals such as Google Maps. The index contains missing when there isn’t enough data to ensure anonymity, and thus the sample sizes for each place can vary.

The mobility index at the following six places were contained:

a. Retail and recreation: restaurants, cafes, shopping centres, theme parks, museums, libraries, and movie theatres.
b. Supermarkets and pharmacies: grocery markets, food warehouses, farmers markets, specialty food shops, drug stores, and pharmacies.
c. Parks: national parks, public beaches, marinas, dog parks, plazas, and public gardens.
d. Public transport: public transport hubs such as subway, bus, and train stations.
e. Workplaces
f. Residential area

#### Weather

The per-day weather conditions were obtained from the Japan Meteorological Agency[2]. This included mean temperature, total precipitation, sunshine duration, total snowfall, and mean wind speed. Data for each prefectural capital (where the Automated Meteorological Data Acquisition System is located) were used in this study. For Saitama prefecture and Shiga prefecture, data in Kumagaya city and Hikone city were used since the system was not equipped in their prefectural capitals. When the data were not recorded for some reasons (e.g. the system was not working properly), the information was imputed from the system of the closest location in the same prefecture.

#### COVID-19 cases

The number of confirmed cases of COVID-19 infections each day for each prefecture were obtained from the Japan Broadcasting Corporation.[3]

#### Stringency index

The stringency index of the Japanese government responses to the pandemic was obtained from the COVID-19 Government Response Tracker published by the research team of the University of Oxford.[4] The index is composed of indicators for school closing, workplace closing, cancellation of public events, restrictions on gathering size, closure of public transport, stay-at-home requirements, restrictions on internal movement, restrictions on international travel, and public information campaign.

#### Vaccination rate

The data on the number of those vaccinated (at least once) by prefecture were obtained from the web site of the Cabinet Secretariat.[5] The data were transformed into the proportions among total population of each prefecture, using the population data mentioned below.

#### Consumption

Consumption data were obtained from the Statistics Bureau of Japan.[6] Monthly average consumptions of households with two or more members up to July 2021 were available.

#### Consumer price index

To obtain the relative changes in consumption from the averages of the past three year, the consumption for each item during the periods (January 2017 - July 2021) were adjusted by the consumer price index. The consumer price index for each consumption item was obtained from the Statistics Bureau of Japan[7].

#### Population

The latest population data (2019) for each prefecture were obtained from the Statistics Bureau of Japan.[8]

#### National holidays

In the study periods, the following dates were coded as national holidays:

Year 2020

11 February: National Foundation Day

23 February: Emperor’s Birthday

24 February: Substitute holiday for Emperor’s Birthday

20 March: Vernal Equinox Day

29 April: Showa Day

3 May: Constitution Memorial Day

4 May: Greenery Day

5 May: Children’s Day

6 May: Substitute holiday for Constitution Memorial Day

23 July: Marine Day

24 July: Sports Day

10 August: Mountain Day

13-16 August: Obon (Japanese Festival of the Dead); While these days are not official national holidays, many Japanese people take summer breaks during this season.

21 September: Respect for the Aged Day

22 September: Autumnal Equinox Day

3 November: Culture Day

23 November: Labour Thanksgiving Day

29-31 December: New Year’s holidays: While these days are not official national holidays, many Japanese people take breaks during this season.

Year 2021

1 January: New Year’s Day

2-3 January: New Year’s holidays: While these days are not official national holidays, many Japanese people take breaks during this season.

11 January: Coming of Age Day

11 February: National Foundation Day

23 February: Emperor’s Birthday

20 March: Vernal Equinox Day

29 April: Showa Day

3 May: Constitution Memorial Day

4 May: Greenery Day

5 May: Children’s Day

22 July: Marine Day

23 July: Sports Day

8 August: Mountain Day

9 August: Substitute holiday for Mountain Day

13-16 August: Obon (Japanese Festival of the Dead); While these days are not official national holidays, many Japanese people take summer breaks during this season.

20 September: Respect for the Aged Day

23 September: Autumnal Equinox Day

---

**Appendix Figure 1.**
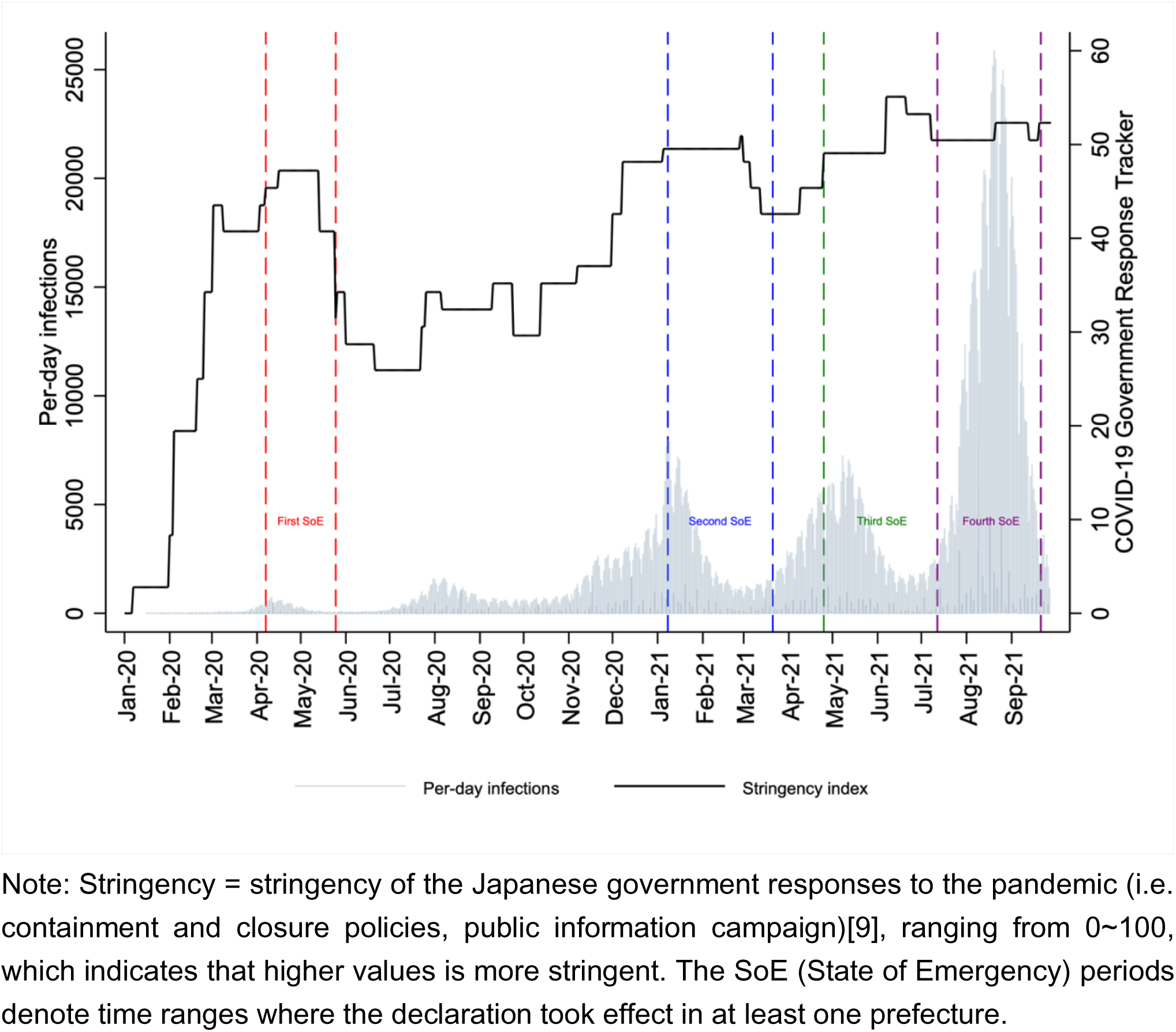
The number of confirmed COVID-19 infections, the state of emergency, and stringency index of the government responses to the pandemic

**Appendix Figure 2.**
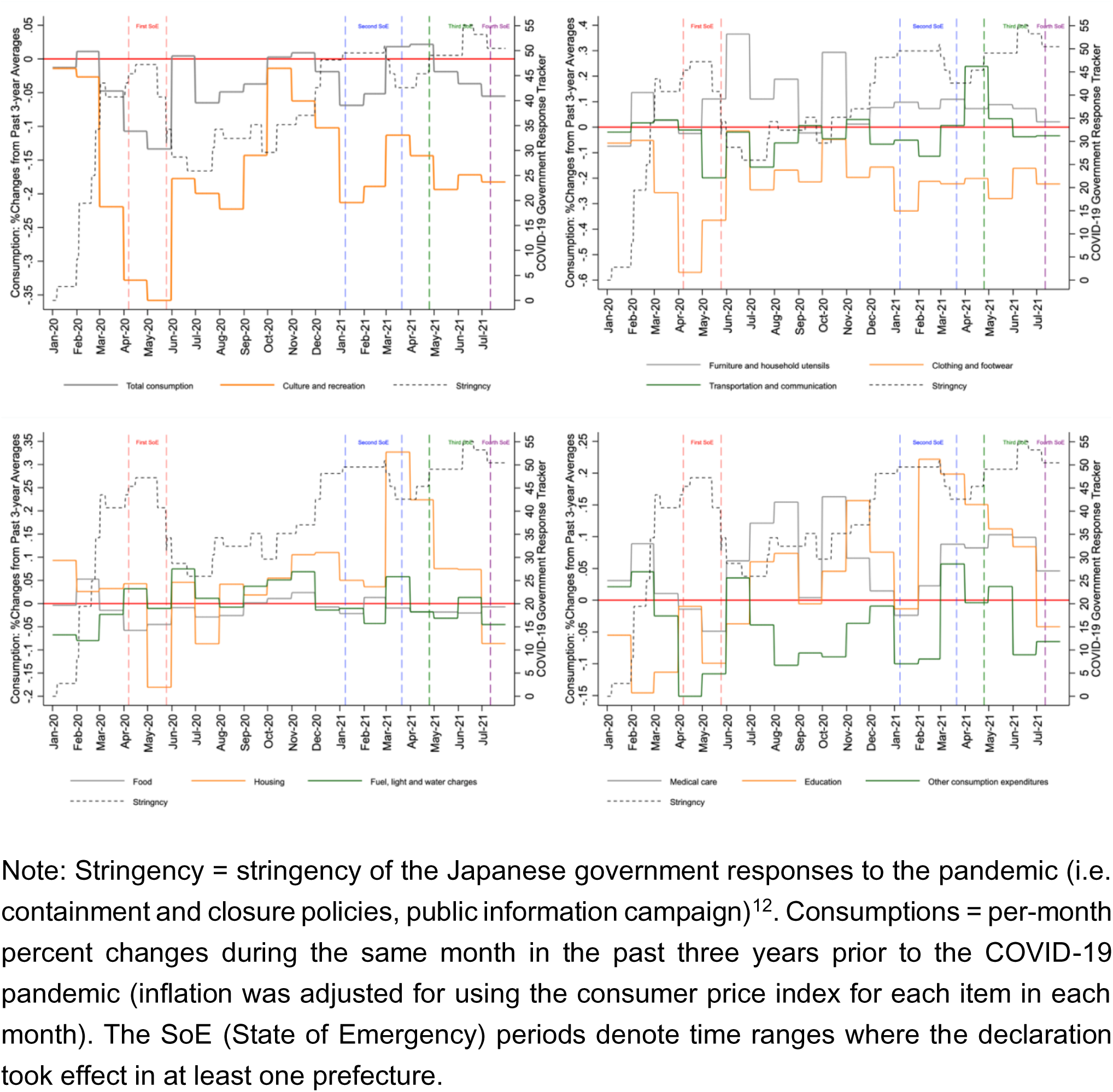
Stringency and consumption

**Appendix Table 1.** SoE and human mobility: Grocery and pharmacies and parks

| Model | Supermarkets and pharmacies |  |  | Parks |  |  |
| --- | --- | --- | --- | --- | --- | --- |
|  | (1) | (2) | (3) | (1) | (2) | (3) |
| Since observation |  | 0.08** | 0.02** |  | 0.00 | -0.01 |
| (day) |  | (0.08 - 0.09) | (0.02 - 0.03) |  | (-0.02 - 0.03) | (-0.04 - 0.02) |
| Since observation^2 |  |  | 0.00** |  |  | 0.00 |
| (day) |  |  | (0.00 - 0.00) |  |  | (-0.00 - 0.00) |
| SoE | -1.83** | -0.81** | -0.54* | -7.54** | -6.01** | -4.63** |
|  | (-2.37 - -1.29) | (-1.31 - -0.32) | (-1.04 - -0.05) | (-9.37 - -5.72) | (-7.64 - -4.39) | (-6.18 - -3.09) |
| SoE days |  | -0.11** | -0.46** |  | -0.17** | -0.79** |
|  |  | (-0.14 - -0.09) | (-0.52 - -0.40) |  | (-0.25 - -0.10) | (-1.10 - -0.49) |
| SoE days^2 |  |  | 0.01** |  |  | 0.02** |
|  |  |  | (0.01 - 0.01) |  |  | (0.01 - 0.02) |
| Covid cases | 0.32** | 0.29** | -0.02# | 0.23** | 0.20** | 0.12* |
| (Last week: 100) | (0.17 - 0.47) | (0.17 - 0.41) | (-0.04 - 0.00) | (0.15 - 0.30) | (0.15 - 0.25) | (0.03 - 0.21) |
| Observations | 27,495 | 27,495 | 27,495 | 27,126 | 27,126 | 27,126 |
| R-squared | 0.45 | 0.47 | 0.55 | 0.62 | 0.62 | 0.62 |
| AIC | 161,396 | 160,374 | 155,857 | 216,147 | 216,077 | 215,949 |

Note: The table presents coefficients with confidence intervals estimated from robust standard errors in parentheses. \*\* p<0.01, \* p<0.05. All models include, prefecture-level fixed-effects, prefecture-by-holiday fixed-effects, prefecture-by-weekdays fixed-effects, prefecture-by-month fixed-effects, and prefecture-by-year fixed-effects, and are weighted by population size. Constants are not presented.
SoE = State of Emergency

**Appendix Table 2.** SoE and human mobility: Public transport and workplaces

| Model | Public transport |  |  | Workplaces |  |  |
| --- | --- | --- | --- | --- | --- | --- |
|  | (1) | (2) | (3) | (1) | (2) | (3) |
| Since observation |  | 0.07** | 0.03** |  | 0.01** | -0.04** |
| (day) |  | (0.05 - 0.09) | (0.01 - 0.06) |  | (0.01 - 0.02) | (-0.05 - -0.02) |
| Since observation^2 |  |  | 0.00** |  |  | 0.00** |
| (day) |  |  | (0.00 - 0.00) |  |  | (0.00 - 0.00) |
| SoE | -15.31** | -12.59** | -10.33** | -6.72** | -4.41** | -3.48** |
|  | (-16.65 - -13.96) | (-14.04 - -11.13) | (-11.82 - -8.83) | (-7.98 - -5.47) | (-5.58 - -3.23) | (-4.56 - -2.40) |
| SoE days |  | -0.31** | -1.37** |  | -0.26** | -0.84** |
|  |  | (-0.38 - -0.23) | (-1.49 - -1.25) |  | (-0.33 - -0.20) | (-1.03 - -0.66) |
| SoE days^2 |  |  | 0.03** |  |  | 0.02** |
|  |  |  | (0.03 - 0.03) |  |  | (0.01 - 0.02) |
| Covid cases | 0.29** | 0.24** | 0.04 | 0.18* | 0.15* | -0.12** |
| (Last week: 100) | (0.08 - 0.51) | (0.09 - 0.39) | (-0.03 - 0.10) | (0.02 - 0.35) | (0.03 - 0.26) | (-0.15 - -0.09) |
| Observations | 27,480 | 27,480 | 27,480 | 27,495 | 27,495 | 27,495 |
| R-squared | 0.71 | 0.72 | 0.74 | 0.80 | 0.81 | 0.82 |
| AIC | 186,108 | 185,094 | 183,478 | 179,086 | 178,339 | 176,558 |

Note) The table presents coefficients with confidence intervals estimated from robust standard errors in parentheses. \*\* p<0.01, \* p<0.05. All models include, prefecture-level fixed-effects, prefecture-by-holiday fixed-effects, prefecture-by-weekdays fixed-effects, prefecture-by-month fixed-effects, and prefecture-by-year fixed-effects, and are weighted by population size. Constants are not presented.
SoE = State of Emergency

**Appendix Table 3.** 1^st^, 2^nd^, 3^rd^, and 4^th^ SoE and human mobility: Grocery and pharmacies and parks

| Model | Supermarkets and pharmacies |  |  | Parks |  |  |
| --- | --- | --- | --- | --- | --- | --- |
|  | (1) | (2) | (3) | (1) | (2) | (3) |
| Since observation |  | 0.01** | -0.02** |  | -0.02** | -0.02** |
| (day) |  | (0.00 - 0.02) | (-0.03 - -0.02) |  | (-0.02 - -0.02) | (-0.02 - -0.02) |
| Since observation^2 |  |  | 0.00** |  |  | 0.00 |
| (day) |  |  | (0.00 - 0.00) |  |  | (-0.00 - 0.00) |
| 1st SoE | -3.45** | -2.36** | -2.26** | -10.58** | -10.06** | -7.26** |
|  | (-3.78 - -3.12) | (-2.96 - -1.76) | (-2.79 - -1.73) | (-13.33 - -7.83) | (-13.51 - -6.62) | (-9.08 - -5.45) |
| 1st SoE (days) |  | -0.06** | -0.36** |  | -0.04 | -1.14** |
|  |  | (-0.07 - -0.04) | (-0.41 - -0.31) |  | (-0.09 - 0.01) | (-1.42 - -0.87) |
| 1st SoE^2 (days) |  |  | 0.01** |  |  | 0.03** |
|  |  |  | (0.01 - 0.01) |  |  | (0.02 - 0.04) |
| 2nd SoE | -4.27** | -0.21 | 2.30** | -7.98** | -0.26 | -0.53 |
|  | (-5.38 - -3.15) | (-0.84 - 0.43) | (0.94 - 3.66) | (-11.31 - -4.65) | (-1.69 - 1.18) | (-2.84 - 1.79) |
| 2nd SoE (days) |  | -0.11** | -0.16** |  | -0.21** | -0.19 |
|  |  | (-0.13 - -0.10) | (-0.24 - -0.08) |  | (-0.26 - -0.16) | (-0.39 - 0.01) |
| 2nd SoE^2 (days) |  |  | 0.00** |  |  | -0.00 |
|  |  |  | (0.00 - 0.00) |  |  | (-0.00 - 0.00) |
| 3rd SoE | 1.51** | -1.41** | -1.86** | -3.02** | -8.29** | -10.84** |
|  | (0.70 - 2.33) | (-2.46 - -0.36) | (-2.72 - -0.99) | (-4.40 - -1.63) | (-10.00 - -6.58) | (-12.85 - -8.82) |
| 3rd SoE (days) |  | 0.10** | 0.18** |  | 0.19** | 0.44** |
|  |  | (0.06 - 0.14) | (0.13 - 0.23) |  | (0.16 - 0.23) | (0.34 - 0.53) |
| 3rd SoE^2 (days) |  |  | -0.00** |  |  | -0.00** |
|  |  |  | (-0.00 - -0.00) |  |  | (-0.00 - -0.00) |
| 4th SoE | 3.40** | 2.51** | -1.80** | -1.53 | -6.59** | -5.55* |
|  | (2.36 - 4.45) | (1.66 - 3.35) | (-2.90 - -0.69) | (-5.88 - 2.81) | (-11.16 - -2.02) | (-10.85 - -0.25) |
| 4th SoE (days) |  | 0.04 | -0.02 |  | 0.29** | -0.07 |
|  |  | (-0.01 - 0.09) | (-0.09 - 0.06) |  | (0.21 - 0.37) | (-0.39 - 0.25) |
| 4th SoE^2 (days) |  |  | 0.00 |  |  | 0.01** |
|  |  |  | (-0.00 - 0.00) |  |  | (0.00 - 0.02) |
| Covid cases | 0.02 | 0.01 | -0.04 | 0.04** | 0.05** | 0.06** |
| (Last week: 100) | (-0.04 - 0.07) | (-0.03 - 0.05) | (-0.10 - 0.02) | (0.03 - 0.06) | (0.03 - 0.06) | (0.03 - 0.09) |
| Observations | 27,495 | 27,495 | 27,495 | 27,126 | 27,126 | 27,126 |
| R-squared | 0.47 | 0.49 | 0.60 | 0.62 | 0.62 | 0.65 |
| AIC | 160,279 | 159,188 | 153,745 | 215,982 | 215,780 | 221,775 |

Note) The table presents coefficients with confidence intervals estimated from robust standard errors in parentheses. \*\* p<0.01, \* p<0.05. All models include prefecture-level fixed-effects, prefecture-by-holiday fixed-effects, prefecture-by-weekdays fixed-effects, prefecture-by-month fixed-effects, and prefecture-by-year fixed-effects, and are weighted by population size. Constants are not presented. SoE = State of Emergency

**Appendix Table 4.** 1^st^, 2^nd^, 3^rd^, and 4^th^ SoE and human mobility: Public transport and workplaces

| Model | Public transport |  |  | Workplaces |  |  |
| --- | --- | --- | --- | --- | --- | --- |
|  | (1) | (2) | (3) | (1) | (2) | (3) |
| Since observation |  | 0.07** | 0.09** |  | 0.01** | -0.02** |
| (day) |  | (0.05 - 0.09) | (0.07 - 0.11) |  | (0.00 - 0.02) | (-0.03 - -0.02) |
| Since observation^2 |  |  | 0.00* |  |  | 0.00** |
| (day) |  |  | (0.00 - 0.00) |  |  | (0.00 - 0.00) |
| 1st SoE | -21.73** | -24.50** | -19.46** | -10.67** | -10.07** | -6.06** |
|  | (-22.55 - -20.90) | (-25.41 - -23.58) | (-20.64 - -18.29) | (-12.43 - -8.91) | (-12.16 - -7.98) | (-7.21 - -4.91) |
| 1st SoE (days) |  | 0.14** | -0.77** |  | -0.03 | -0.42** |
|  |  | (0.11 - 0.17) | (-0.96 - -0.57) |  | (-0.07 - 0.01) | (-0.51 - -0.32) |
| 1st SoE^2 (days) |  |  | 0.02** |  |  | 0.01** |
|  |  |  | (0.02 - 0.03) |  |  | (0.01 - 0.01) |
| 2nd SoE | -15.92** | -16.64** | -3.31** | -7.97** | -3.19* | 5.89** |
|  | (-17.22 - -14.63) | (-18.91 - -14.37) | (-4.53 - -2.09) | (-9.60 - -6.34) | (-5.72 - -0.67) | (4.86 - 6.93) |
| 2nd SoE (days) |  | 0.02 | -1.10** |  | -0.13** | -0.73** |
|  |  | (-0.07 - 0.11) | (-1.23 - -0.98) |  | (-0.19 - -0.07) | (-0.81 - -0.66) |
| 2nd SoE^2 (days) |  |  | 0.02** |  |  | 0.01** |
|  |  |  | (0.01 - 0.02) |  |  | (0.01 - 0.01) |
| 3rd SoE | -2.74** | -4.40** | -5.21** | 2.37** | 0.24 | 0.04 |
|  | (-4.49 - -1.00) | (-6.95 - -1.85) | (-7.64 - -2.78) | (0.76 - 3.97) | (-2.60 - 3.08) | (-2.80 - 2.87) |
| 3rd SoE (days) |  | 0.08* | 0.12 |  | 0.07* | 0.13 |
|  |  | (0.01 - 0.15) | (-0.01 - 0.26) |  | (0.01 - 0.14) | (-0.03 - 0.29) |
| 3rd SoE^2 (days) |  |  | -0.00 |  |  | -0.00* |
|  |  |  | (-0.00 - 0.00) |  |  | (-0.00 - -0.00) |
| 4th SoE | -5.78** | -5.96** | -9.16** | -0.79 | -0.03 | -3.46** |
|  | (-8.45 - -3.12) | (-9.56 - -2.37) | (-12.04 - -6.28) | (-1.89 - 0.31) | (-0.75 - 0.70) | (-4.15 - -2.77) |
| 4th SoE (days) |  | -0.00 | -0.04 |  | -0.05** | -0.13* |
|  |  | (-0.08 - 0.07) | (-0.16 - 0.07) |  | (-0.08 - -0.01) | (-0.22 - -0.03) |
| 4th SoE^2 (days) |  |  | 0.00 |  |  | 0.00 |
|  |  |  | (-0.00 - 0.00) |  |  | (-0.00 - 0.00) |
| Covid cases | 0.03 | 0.04 | -0.04 | 0.02 | 0.01 | -0.04 |
| (Last week: 100) | (-0.03 - 0.09) | (-0.01 - 0.10) | (-0.15 - 0.07) | (-0.04 - 0.07) | (-0.03 - 0.05) | (-0.10 - 0.02) |
| Observations | 27,480 | 27,480 | 27,480 | 27,495 | 27,495 | 27,495 |
| R-squared | 0.74 | 0.74 | 0.71 | 0.81 | 0.82 | 0.81 |
| AIC | 183,032 | 182,711 | 190,113 | 177,216 | 177,021 | 174,688 |

Note) The table presents coefficients with confidence intervals estimated from robust standard errors in parentheses. \*\* p<0.01, \* p<0.05. All models include prefecture-level fixed-effects, prefecture-by-holiday fixed-effects, prefecture-by-weekdays fixed-effects, prefecture-by-month fixed-effects, and prefecture-by-year fixed-effects, and are weighted by population size. Constants are not presented. SoE = State of Emergency

**Appendix Table 5.**
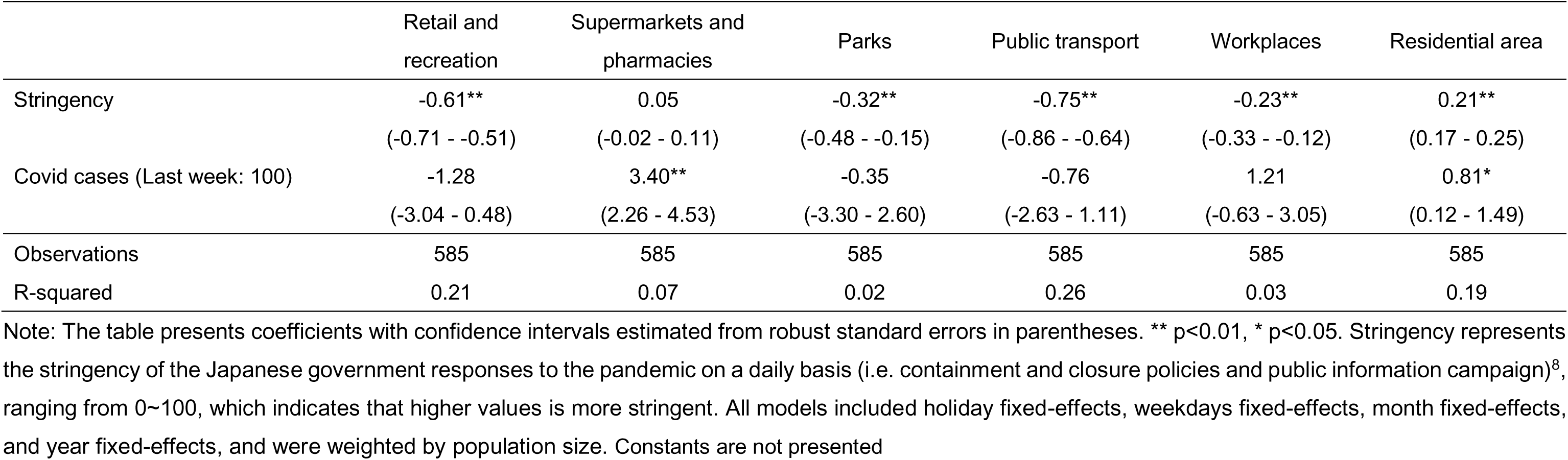
Stringency index and mobility

**Appendix Table 6.** Containment and closure policies and mobility

|  | Retail and recreation | Supermarkets and pharmacies | Parks | Public transport | Workplaces | Residential area |
| --- | --- | --- | --- | --- | --- | --- |
| School closing | -6.47**<br>(-11.19 - -1.74) | -5.71**<br>(-8.62 - -2.80) | 4.69<br>(-3.86 - 13.25) | -13.11**<br>(-17.82 - -8.40) | -9.34**<br>(-14.39 - -4.30) | 3.27**<br>(1.38 - 5.17) |
| Workplace closing | -4.56**<br>(-6.54 - -2.58) | 1.68**<br>(0.46 - 2.90) | -5.52**<br>(-9.12 - -1.93) | -4.35**<br>(-6.33 - -2.38) | 1.17<br>(-0.95 - 3.29) | 1.28**<br>(0.49 - 2.08) |
| Cancel public events | 0.12<br>(-5.69 - 5.93) | 1.24<br>(-2.34 - 4.81) | 1.86<br>(-8.66 - 12.39) | -1.53<br>(-7.32 - 4.26) | 1.18<br>(-5.02 - 7.38) | -0.48<br>(-2.81 - 1.85) |
| Restrictions on gatherings | 10.69**<br>(4.97 - 16.41) | 14.39**<br>(10.87 - 17.91) | 8.01<br>(-2.35 - 18.37) | 1.33<br>(-4.37 - 7.03) | 12.25**<br>(6.14 - 18.36) | -2.91*<br>(-5.21 - -0.62) |
| Close public transport | -9.01**<br>(-12.47 - -5.55) | -3.68**<br>(-5.81 - -1.55) | -14.52**<br>(-20.79 - -8.25) | -11.38**<br>(-14.83 - -7.93) | -4.11*<br>(-7.80 - -0.42) | 2.86**<br>(1.48 - 4.25) |
| Stay-at-home orders | -9.36**<br>(-11.70 - -7.03) | -5.09**<br>(-6.53 - -3.65) | -3.19<br>(-7.42 - 1.04) | -9.66**<br>(-11.99 - -7.33) | -7.79**<br>(-10.29 - -5.30) | 3.02**<br>(2.08 - 3.96) |
| Restrictions on internal movement | 0.34<br>(-2.25 - 2.93) | 2.19**<br>(0.59 - 3.78) | -0.87<br>(-5.57 - 3.83) | -1.41<br>(-4.00 - 1.17) | 0.37<br>(-2.40 - 3.14) | 0.25<br>(-0.79 - 1.29) |
| Covid cases (Last week) | -9.40**<br>(-11.19 - -7.62) | 0.74<br>(-0.35 - 1.84) | -5.11**<br>(-8.34 - -1.88) | -10.50**<br>(-12.28 - -8.72) | -4.05**<br>(-5.95 - -2.14) | 3.58**<br>(2.87 - 4.30) |
| Observations | 585 | 585 | 585 | 585 | 585 | 585 |
| R-squared | 0.21 | 0.37 | 0.07 | 0.32 | 0.02 | 0.09 |

Note: The table presents coefficients with confidence intervals estimated from robust standard errors in parentheses. \*\* $p < 0.01$ , \* $p < 0.05$ . Each of the national Japanese government responses to the pandemic on a daily basis was obtained from the research team of the University of Oxford<sup>8</sup>. As international travel controls and public information have been persistent during the pandemic, these variables were omitted from the estimations by the fixed-effects models. All models included holiday fixed-effects, weekdays fixed-effects, month fixed-effects, and year fixed-effects, and were weighted by population size. Constants are not presented.

**Appendix Table 7.** Quasi-SoE, SoE, and human mobility: Retail and recreation, grocery and pharmacies, and parks

|  | Retail and recreation |  | Supermarkets and pharmacies |  | Parks |  |
| --- | --- | --- | --- | --- | --- | --- |
| Model | (1) | (2) | (1) | (2) | (1) | (2) |
| Since observation (day) |  | 0.11**<br>(0.10 - 0.12) |  | 0.08**<br>(0.08 - 0.08) |  | 0.01<br>(-0.02 - 0.04) |
| Quasi-SoE | -2.80**<br>(-3.69 - -1.91) | 0.46<br>(-1.01 - 1.93) | 2.38**<br>(1.42 - 3.34) | 1.13**<br>(0.53 - 1.72) | -2.90**<br>(-4.75 - -1.05) | -2.51**<br>(-4.33 - -0.69) |
| Quasi-SoE days |  | -0.08**<br>(-0.09 - -0.06) |  | 0.05**<br>(0.04 - 0.06) |  | 0.01<br>(-0.03 - 0.05) |
| SoE | -13.56**<br>(-14.39 - -12.72) | -10.70**<br>(-11.76 - -9.63) | -1.25**<br>(-1.74 - -0.77) | 0.07<br>(-0.41 - 0.56) | -8.25**<br>(-10.15 - -6.35) | -6.75**<br>(-8.57 - -4.93) |
| SoE days |  | -0.28**<br>(-0.36 - -0.21) |  | -0.15**<br>(-0.16 - -0.13) |  | -0.16**<br>(-0.24 - -0.07) |
| Covid cases<br>(Last week: 100) | 0.07<br>(-0.05 - 0.19) | 0.01<br>(-0.04 - 0.07) | 0.33**<br>(0.17 - 0.49) | 0.30**<br>(0.17 - 0.43) | 0.21**<br>(0.15 - 0.28) | 0.19**<br>(0.15 - 0.23) |
| Observations | 27,495 | 27,495 | 27,495 | 27,495 | 27,126 | 27,126 |
| R-squared | 0.76 | 0.77 | 0.45 | 0.48 | 0.62 | 0.62 |

Note: The table presents coefficients with confidence intervals estimated from robust standard errors in parentheses. \*\* p<0.01, \* p<0.05. All models included prefecture-level fixed-effects, prefecture-by-holiday fixed-effects, prefecture-by-weekdays fixed-effects, prefecture-by-month fixed-effects, and prefecture-by-year fixed-effects, and were weighted by population size. Constants are not presented.
SoE = State of Emergency

**Appendix Table 8.** Quasi-SoE, SoE, and human mobility: Public transport, workplaces, and residential area

| Model | Public transport |  | Workplaces |  | Residential area |  |
| --- | --- | --- | --- | --- | --- | --- |
|  | (1) | (2) | (1) | (2) | (1) | (2) |
| Since observation (day) |  | 0.07**<br>(0.06 - 0.09) |  | 0.01**<br>(0.00 - 0.02) |  | -0.02**<br>(-0.02 - -0.02) |
| Quasi-SoE | -3.06**<br>(-4.24 - -1.89) | 0.00<br>(-1.55 - 1.56) | 1.53**<br>(0.50 - 2.57) | 2.73**<br>(1.53 - 3.93) | 0.65**<br>(0.34 - 0.96) | -0.48*<br>(-0.89 - -0.06) |
| Quasi-SoE days |  | -0.07**<br>(-0.09 - -0.05) |  | -0.00<br>(-0.02 - 0.02) |  | 0.02**<br>(0.01 - 0.03) |
| SoE | -16.05**<br>(-17.38 - -14.72) | -13.25**<br>(-15.05 - -11.44) | -6.35**<br>(-7.46 - -5.24) | -3.57**<br>(-4.60 - -2.54) | 4.61**<br>(4.21 - 5.01) | 3.44**<br>(2.96 - 3.91) |
| SoE days |  | -0.28**<br>(-0.37 - -0.19) |  | -0.29**<br>(-0.36 - -0.22) |  | 0.12**<br>(0.08 - 0.16) |
| Covid cases<br>(Last week: 100) | 0.28**<br>(0.07 - 0.48) | 0.23**<br>(0.09 - 0.36) | 0.19*<br>(0.02 - 0.36) | 0.16*<br>(0.03 - 0.28) | -0.01<br>(-0.05 - 0.03) | 0.01<br>(-0.01 - 0.03) |
| Observations | 27,480 | 27,480 | 27,495 | 27,495 | 27,495 | 27,495 |
| R-squared | 0.71 | 0.72 | 0.80 | 0.81 | 0.80 | 0.81 |

Note: The table presents coefficients with confidence intervals estimated from robust standard errors in parentheses. \*\* p<0.01, \* p<0.05. All models included prefecture-level fixed-effects, prefecture-by-holiday fixed-effects, prefecture-by-weekdays fixed-effects, prefecture-by-month fixed-effects, and prefecture-by-year fixed-effects, and were weighted by population size. Constants are not presented.
SoE = State of Emergency

**Appendix Table 9.** Quasi-SoE, 1st, 2nd, 3rd, and 4th SoE, and human mobility: Retail and recreation, grocery and pharmacies, and parks

| Model | Retail and recreation |  | Supermarkets and pharmacies |  | Parks |  |
| --- | --- | --- | --- | --- | --- | --- |
|  | (1) | (2) | (1) | (2) | (1) | (2) |
| Quasi-SoE | 1.06<br>(-0.56 - 2.69) | 2.50**<br>(0.71 - 4.29) | 4.23**<br>(3.33 - 5.13) | 1.97**<br>(1.26 - 2.68) | -1.07<br>(-3.12 - 0.99) | -2.03*<br>(-3.91 - -0.15) |
| Quasi-SoE days |  | -0.06**<br>(-0.07 - -0.04) |  | 0.06**<br>(0.05 - 0.08) |  | 0.02<br>(-0.03 - 0.07) |
| 1st SoE | -19.38**<br>(-20.36 - -18.40) | -20.35**<br>(-21.51 - -19.18) | -3.36**<br>(-3.77 - -2.96) | -3.08**<br>(-3.51 - -2.65) | -10.60**<br>(-13.34 - -7.86) | -10.05**<br>(-13.70 - -6.39) |
| 1st SoE (days) |  | 0.07**<br>(0.03 - 0.10) |  | -0.04**<br>(-0.05 - -0.03) |  | -0.05<br>(-0.10 - 0.00) |
| 2nd SoE | -10.48**<br>(-12.28 - -8.67) | -8.90**<br>(-9.88 - -7.93) | -2.83**<br>(-4.04 - -1.63) | 0.34<br>(-0.50 - 1.19) | -8.35**<br>(-12.02 - -4.67) | -0.43<br>(-2.03 - 1.18) |
| 2nd SoE (days) |  | -0.05**<br>(-0.09 - -0.02) |  | -0.08**<br>(-0.11 - -0.06) |  | -0.22**<br>(-0.27 - -0.16) |
| 3rd SoE | -2.72*<br>(-5.38 - -0.07) | -5.75**<br>(-7.98 - -3.52) | 3.64**<br>(2.68 - 4.61) | 0.15<br>(-1.07 - 1.36) | -3.55**<br>(-5.60 - -1.51) | -9.09**<br>(-11.30 - -6.88) |
| 3rd SoE (days) |  | 0.12**<br>(0.08 - 0.16) |  | 0.11**<br>(0.06 - 0.16) |  | 0.19**<br>(0.16 - 0.23) |
| 4th SoE | -4.05**<br>(-6.51 - -1.59) | -5.20**<br>(-7.68 - -2.71) | 5.61**<br>(4.00 - 7.22) | 4.68**<br>(3.30 - 6.05) | -2.09<br>(-6.41 - 2.23) | -7.24**<br>(-11.95 - -2.52) |
| 4th SoE (days) |  | 0.02<br>(-0.03 - 0.06) |  | 0.06<br>(-0.01 - 0.12) |  | 0.29**<br>(0.21 - 0.37) |
| Observations | 27,495 | 27,495 | 27,495 | 27,495 | 27,126 | 27,126 |
| R-squared | 0.78 | 0.79 | 0.48 | 0.51 | 0.62 | 0.62 |

Note: The table presents coefficients with confidence intervals estimated from robust standard errors in parentheses. \*\* p<0.01, \* p<0.05. All models included the per-day confirmed infections in the past seven days, prefecture-level fixed-effects, prefecture-by-holiday fixed-effects, prefecture-by-weekdays fixed-effects, prefecture-by-month fixed-effects, and prefecture-by-year fixed-effects, and were weighted by population size. In addition, days since the start of the observation is included in Model (2).

**Appendix Table 10.** Quasi-SoE, 1st, 2nd, 3rd, and 4th SoE and human mobility: Public transport, workplaces, and residential area

| Model | Public transport |  | Workplaces |  | Residential area |  |
| --- | --- | --- | --- | --- | --- | --- |
|  | (1) | (2) | (1) | (2) | (1) | (2) |
| Quasi-SoE | 1.35<br>(-0.67 - 3.36) | 3.10**<br>(1.06 - 5.13) | 4.96**<br>(3.57 - 6.35) | 4.83**<br>(3.21 - 6.46) | -0.81*<br>(-1.44 - -0.18) | -1.37**<br>(-2.01 - -0.72) |
| Quasi-SoE days |  | -0.06**<br>(-0.07 - -0.04) |  | -0.00<br>(-0.02 - 0.01) |  | 0.02**<br>(0.02 - 0.03) |
| 1st SoE | -21.70**<br>(-22.57 - -20.83) | -24.28**<br>(-25.31 - -23.26) | -10.56**<br>(-12.24 - -8.88) | -10.46**<br>(-12.59 - -8.33) | 6.62**<br>(5.91 - 7.33) | 6.77**<br>(6.02 - 7.52) |
| 1st SoE (days) |  | 0.15**<br>(0.13 - 0.17) |  | -0.01<br>(-0.04 - 0.03) |  | -0.02**<br>(-0.03 - -0.01) |
| 2nd SoE | -15.47**<br>(-16.99 - -13.95) | -16.51**<br>(-18.76 - -14.26) | -6.29**<br>(-7.68 - -4.90) | -2.59*<br>(-4.85 - -0.33) | 3.82**<br>(3.12 - 4.52) | 2.55**<br>(1.07 - 4.03) |
| 2nd SoE (days) |  | 0.02<br>(-0.07 - 0.11) |  | -0.11**<br>(-0.17 - -0.04) |  | 0.04*<br>(0.00 - 0.08) |
| 3rd SoE | -2.06<br>(-4.69 - 0.56) | -3.45*<br>(-6.53 - -0.37) | 4.87**<br>(3.10 - 6.64) | 2.54<br>(-0.02 - 5.10) | -0.17<br>(-1.12 - 0.77) | 0.11<br>(-0.86 - 1.07) |
| 3rd SoE (days) |  | 0.08*<br>(0.00 - 0.15) |  | 0.08*<br>(0.01 - 0.15) |  | -0.01<br>(-0.03 - 0.01) |
| 4th SoE | -5.08**<br>(-8.48 - -1.68) | -5.52*<br>(-9.65 - -1.38) | 1.80*<br>(0.02 - 3.59) | 2.28**<br>(0.65 - 3.90) | 1.57*<br>(0.38 - 2.75) | 1.47**<br>(0.39 - 2.56) |
| 4th SoE (days) |  | -0.01<br>(-0.08 - 0.06) |  | -0.04**<br>(-0.06 - -0.02) |  | 0.02<br>(-0.01 - 0.05) |
| Observations | 27,480 | 27,480 | 27,495 | 27,495 | 27,495 | 27,495 |
| R-squared | 0.74 | 0.75 | 0.82 | 0.82 | 0.82 | 0.82 |

Note: The table presents coefficients with confidence intervals estimated from robust standard errors in parentheses. \*\* p<0.01, \* p<0.05. All models included the per-day confirmed infections in the past seven days, prefecture-level fixed-effects, prefecture-by-holiday fixed-effects, prefecture-by-weekdays fixed-effects, prefecture-by-month fixed-effects, and prefecture-by-year fixed-effects, and were weighted by population size. In addition, days since the start of the observation is included in Model (2).

**Appendix Table 11.** SoE and human mobility for retail and recreation, grocery and pharmacies, and parks: Comparison across regions by COVID-19 confirmed cases

|  | Retail and recreation |  |  | Supermarkets and pharmacies |  |  | Parks |  |  |
| --- | --- | --- | --- | --- | --- | --- | --- | --- | --- |
|  | High |  |  |  |  |  |  |  |  |
| Model | (1) | (2) | (3) | (1) | (2) | (3) | (1) | (2) | (3) |
| SoE | -12.44** | -9.76** | -7.72** | -1.63** | -0.72* | -0.51 | -6.44** | -5.33** | -4.54** |
|  | (-13.37 - -11.50) | (-10.56 - -8.96) | (-8.53 - -6.92) | (-2.24 - -1.03) | (-1.27 - -0.18) | (-1.05 - 0.03) | (-8.15 - -4.73) | (-6.99 - -3.67) | (-6.15 - -2.93) |
| SoE days |  | -0.33** | -1.38** |  | -0.11** | -0.46** |  | -0.14** | -0.60** |
|  |  | (-0.39 - -0.26) | (-1.55 - -1.20) |  | (-0.13 - -0.09) | (-0.53 - -0.40) |  | (-0.20 - -0.07) | (-0.89 - -0.31) |
| SoE days^2 |  |  | 0.03** |  |  | 0.01** |  |  | 0.01** |
|  |  |  | (0.02 - 0.03) |  |  | (0.01 - 0.01) |  |  | (0.01 - 0.02) |
| Observations | 13,455 | 13,455 | 13,455 | 13,455 | 13,455 | 13,455 | 13,436 | 13,436 | 13,436 |
| R-squared | 0.75 | 0.77 | 0.79 | 0.41 | 0.43 | 0.52 | 0.57 | 0.57 | 0.57 |
|  | Low |  |  |  |  |  |  |  |  |
| SoE | -17.48** | -19.96** | -11.17** | -3.82** | -2.42** | -0.52 | -19.09** | -16.03** | -6.70** |
|  | (-18.69 - -16.28) | (-21.36 - -18.55) | (-12.62 - -9.72) | (-4.41 - -3.22) | (-3.20 - -1.64) | (-1.19 - 0.15) | (-21.49 - -16.70) | (-18.65 - -13.42) | (-10.19 - -3.20) |
| SoE days |  | 0.17** | -1.61** |  | -0.09** | -0.76** |  | -0.20** | -1.98** |
|  |  | (0.14 - 0.20) | (-1.77 - -1.45) |  | (-0.11 - -0.07) | (-0.84 - -0.67) |  | (-0.32 - -0.09) | (-2.50 - -1.47) |
| SoE days^2 |  |  | 0.06** |  |  | 0.02** |  |  | 0.06** |
|  |  |  | (0.05 - 0.06) |  |  | (0.02 - 0.03) |  |  | (0.05 - 0.07) |
| Observations | 14,040 | 14,040 | 14,040 | 14,040 | 14,040 | 14,040 | 13,690 | 13,690 | 13,690 |
| R-squared | 0.66 | 0.67 | 0.68 | 0.54 | 0.55 | 0.61 | 0.64 | 0.64 | 0.64 |

Note: The table presents coefficients with confidence intervals estimated from robust standard errors in parentheses. \*\* p<0.01, \* p<0.05. All models included the per-day confirmed infections in the past seven days, prefecture-level fixed-effects, prefecture-by-holiday fixed-effects, prefecture-by-weekdays fixed-effects, prefecture-by-month fixed-effects, and prefecture-by-year fixed-effects, and were weighted by population size. A prefecture was classified as 'High' if the cumulative number of confirmed cases in that area was above the median of all prefectures within the study period; 'Low' prefectures were those with median or below median confirmed cases. SoE = State of Emergency

**Appendix Table 12.**
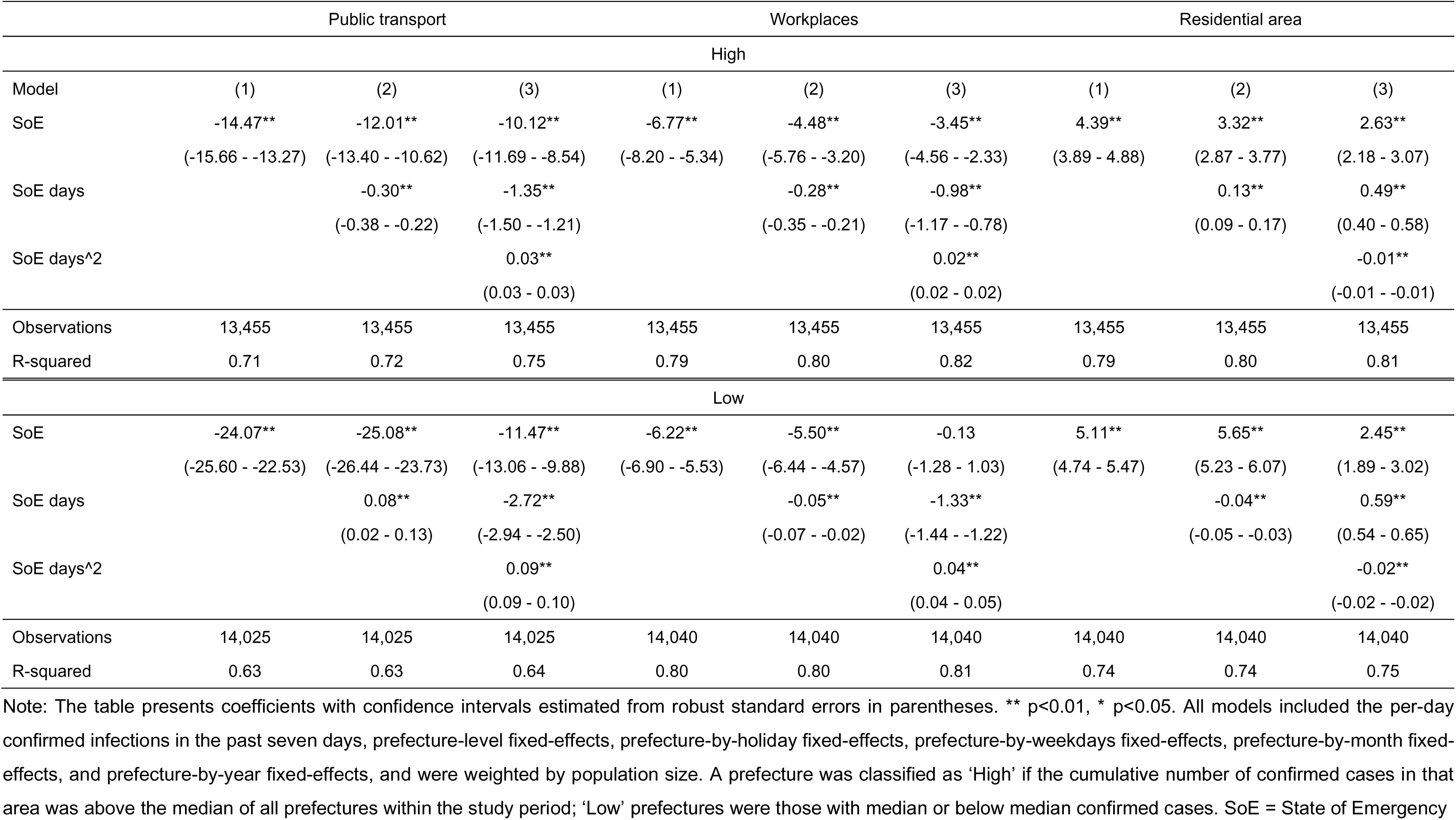
SoE and human mobility for public transport, workplaces, and residential area: Comparison across regions by COVID-19 confirmed cases

|  | Public transport |  |  | Workplaces |  |  | Residential area |  |  |
| --- | --- | --- | --- | --- | --- | --- | --- | --- | --- |
| High |  |  |  |  |  |  |  |  |  |
| Model | (1) | (2) | (3) | (1) | (2) | (3) | (1) | (2) | (3) |
| SoE | -14.47** | -12.01** | -10.12** | -6.77** | -4.48** | -3.45** | 4.39** | 3.32** | 2.63** |
|  | (-15.66 - -13.27) | (-13.40 - -10.62) | (-11.69 - -8.54) | (-8.20 - -5.34) | (-5.76 - -3.20) | (-4.56 - -2.33) | (3.89 - 4.88) | (2.87 - 3.77) | (2.18 - 3.07) |
| SoE days |  | -0.30** | -1.35** |  | -0.28** | -0.98** |  | 0.13** | 0.49** |
|  |  | (-0.38 - -0.22) | (-1.50 - -1.21) |  | (-0.35 - -0.21) | (-1.17 - -0.78) |  | (0.09 - 0.17) | (0.40 - 0.58) |
| SoE days^2 |  |  | 0.03** |  |  | 0.02** |  |  | -0.01** |
|  |  |  | (0.03 - 0.03) |  |  | (0.02 - 0.02) |  |  | (-0.01 - -0.01) |
| Observations | 13,455 | 13,455 | 13,455 | 13,455 | 13,455 | 13,455 | 13,455 | 13,455 | 13,455 |
| R-squared | 0.71 | 0.72 | 0.75 | 0.79 | 0.80 | 0.82 | 0.79 | 0.80 | 0.81 |
| Low |  |  |  |  |  |  |  |  |  |
| SoE | -24.07** | -25.08** | -11.47** | -6.22** | -5.50** | -0.13 | 5.11** | 5.65** | 2.45** |
|  | (-25.60 - -22.53) | (-26.44 - -23.73) | (-13.06 - -9.88) | (-6.90 - -5.53) | (-6.44 - -4.57) | (-1.28 - 1.03) | (4.74 - 5.47) | (5.23 - 6.07) | (1.89 - 3.02) |
| SoE days |  | 0.08** | -2.72** |  | -0.05** | -1.33** |  | -0.04** | 0.59** |
|  |  | (0.02 - 0.13) | (-2.94 - -2.50) |  | (-0.07 - -0.02) | (-1.44 - -1.22) |  | (-0.05 - -0.03) | (0.54 - 0.65) |
| SoE days^2 |  |  | 0.09** |  |  | 0.04** |  |  | -0.02** |
|  |  |  | (0.09 - 0.10) |  |  | (0.04 - 0.05) |  |  | (-0.02 - -0.02) |
| Observations | 14,025 | 14,025 | 14,025 | 14,040 | 14,040 | 14,040 | 14,040 | 14,040 | 14,040 |
| R-squared | 0.63 | 0.63 | 0.64 | 0.80 | 0.80 | 0.81 | 0.74 | 0.74 | 0.75 |

Note: The table presents coefficients with confidence intervals estimated from robust standard errors in parentheses. \*\* p<0.01, \* p<0.05. All models included the per-day confirmed infections in the past seven days, prefecture-level fixed-effects, prefecture-by-holiday fixed-effects, prefecture-by-weekdays fixed-effects, prefecture-by-month fixed-effects, and prefecture-by-year fixed-effects, and were weighted by population size. A prefecture was classified as 'High' if the cumulative number of confirmed cases in that area was above the median of all prefectures within the study period; 'Low' prefectures were those with median or below median confirmed cases. SoE = State of Emergency

**Appendix Table 13.** 1^st^, 2^nd^, and 3^rd^ SoE and human mobility: Retail and recreation, grocery and pharmacies, and parks in prefectures with the high cumulative number of confirmed cases

| Model | Retail and recreation |  |  | Supermarkets and pharmacies |  |  | Parks |  |  |
| --- | --- | --- | --- | --- | --- | --- | --- | --- | --- |
|  | (1) | (2) | (3) | (1) | (2) | (3) | (1) | (2) | (3) |
| 1st SoE | -19.91** | -21.32** | -15.35** | -3.36** | -2.24** | -2.52** | -8.27** | -6.93** | -4.32** |
|  | (-21.05 - -18.77) | (-22.73 - -19.91) | (-17.09 - -13.62) | (-3.74 - -2.98) | (-3.02 - -1.46) | (-3.35 - -1.70) | (-10.90 - -5.64) | (-10.76 - -3.10) | (-6.72 - -1.92) |
| 1st SoE (days) |  | 0.06** | -0.66** |  | -0.06** | -0.33** |  | -0.08** | -1.01** |
|  |  | (0.02 - 0.10) | (-0.86 - -0.45) |  | (-0.08 - -0.04) | (-0.39 - -0.27) |  | (-0.14 - -0.02) | (-1.31 - -0.71) |
| 1st SoE^2 (days) |  |  | 0.02** |  |  | 0.01** |  |  | 0.02** |
|  |  |  | (0.01 - 0.02) |  |  | (0.01 - 0.01) |  |  | (0.01 - 0.03) |
| 2nd SoE | -10.75** | -8.91** | -2.63** | -4.29** | -0.21 | 2.20** | -8.37** | -0.44 | -0.81 |
|  | (-12.77 - -8.73) | (-9.96 - -7.87) | (-4.23 - -1.02) | (-5.46 - -3.11) | (-0.86 - 0.45) | (0.72 - 3.68) | (-11.89 - -4.85) | (-1.95 - 1.06) | (-3.16 - 1.55) |
| 2nd SoE (days) |  | -0.05* | -0.55** |  | -0.11** | -0.12* |  | -0.21** | -0.11 |
|  |  | (-0.09 - -0.01) | (-0.66 - -0.45) |  | (-0.13 - -0.10) | (-0.21 - -0.03) |  | (-0.27 - -0.16) | (-0.31 - 0.08) |
| 2nd SoE^2 (days) |  |  | 0.01** |  |  | 0.00 |  |  | -0.00 |
|  |  |  | (0.01 - 0.01) |  |  | (-0.00 - 0.00) |  |  | (-0.00 - 0.00) |
| 3rd SoE | -3.34** | -6.65** | -7.46** | 1.53** | -1.34* | -1.92** | -2.68** | -7.58** | -10.01** |
|  | (-5.46 - -1.21) | (-8.42 - -4.88) | (-9.03 - -5.90) | (0.67 - 2.38) | (-2.43 - -0.26) | (-2.85 - -0.99) | (-4.12 - -1.25) | (-9.25 - -5.91) | (-12.20 - -7.82) |
| 3rd SoE (days) |  | 0.13** | 0.15** |  | 0.10** | 0.18** |  | 0.17** | 0.39** |
|  |  | (0.09 - 0.17) | (0.08 - 0.23) |  | (0.05 - 0.14) | (0.13 - 0.24) |  | (0.14 - 0.21) | (0.29 - 0.50) |
| 3rd SoE^2 (days) |  |  | -0.00 |  |  | -0.00** |  |  | -0.00** |
|  |  |  | (-0.00 - 0.00) |  |  | (-0.00 - -0.00) |  |  | (-0.00 - -0.00) |
| 4th SoE | -4.54** | -5.24** | -7.50** | 3.42** | 2.50** | -2.42** | -2.10 | -7.02** | -7.54** |
|  | (-6.37 - -2.72) | (-7.22 - -3.25) | (-9.41 - -5.60) | (2.32 - 4.51) | (1.64 - 3.36) | (-3.57 - -1.27) | (-6.45 - 2.24) | (-11.67 - -2.37) | (-12.82 - -2.25) |
| 4th SoE (days) |  | 0.02 | 0.00 |  | 0.04 | -0.04 |  | 0.28** | -0.15 |
|  |  | (-0.02 - 0.07) | (-0.13 - 0.13) |  | (-0.01 - 0.10) | (-0.12 - 0.03) |  | (0.20 - 0.37) | (-0.49 - 0.18) |
| 4th SoE^2 (days) |  |  | 0.00 |  |  | 0.00 |  |  | 0.01** |
|  |  |  | (-0.00 - 0.00) |  |  | (-0.00 - 0.00) |  |  | (0.00 - 0.02) |
| Observations | 13,455 | 13,455 | 13,455 | 13,455 | 13,455 | 13,455 | 13,436 | 13,436 | 13,436 |
| R-squared | 0.78 | 0.79 | 0.75 | 0.44 | 0.46 | 0.54 | 0.57 | 0.58 | 0.58 |

Note: The table presents coefficients with confidence intervals estimated from robust standard errors in parentheses. \*\* $p < 0.01$ , \* $p < 0.05$ . All models included the per-day confirmed infections in the past seven days, prefecture-level fixed-effects, prefecture-by-holiday fixed-effects, prefecture-by-weekdays fixed-effects, prefecture-by-month fixed-effects, and prefecture-by-year fixed-effects, and were weighted by population size. In addition, days since the start of the observation is included in Model (2), and additionally its squared term in Model (3). Full results are available upon request. A prefecture was classified as 'High' if the cumulative number of confirmed cases in that area was above the median of all prefectures within the study period. SoE = State of Emergency

**Appendix Table 14.** 1^st^, 2^nd^, and 3^rd^ SoE and human mobility: Public transport, workplaces, and residential area in prefectures with the high cumulative number of confirmed cases

| Model | Public transport |  |  | Workplaces |  |  | Residential area |  |  |
| --- | --- | --- | --- | --- | --- | --- | --- | --- | --- |
|  | (1) | (2) | (3) | (1) | (2) | (3) | (1) | (2) | (3) |
| 1st SoE | -21.08** | -24.00** | -19.97** | -11.88** | -11.88** | -8.41** | 7.05** | 7.42** | 5.18** |
|  | (-21.89 - -20.28) | (-25.07 - -22.94) | (-21.77 - -18.17) | (-13.76 - -9.99) | (-14.25 - -9.51) | (-9.85 - -6.97) | (6.22 - 7.87) | (6.51 - 8.33) | (4.53 - 5.84) |
| 1st SoE (days) |  | 0.14** | -0.56** |  | -0.00 | -0.40** |  | -0.02** | 0.21** |
|  |  | (0.11 - 0.17) | (-0.73 - -0.39) |  | (-0.04 - 0.04) | (-0.52 - -0.27) |  | (-0.03 - -0.01) | (0.13 - 0.28) |
| 1st SoE^2 (days) |  |  | 0.02** |  |  | 0.01** |  |  | -0.01** |
|  |  |  | (0.01 - 0.02) |  |  | (0.01 - 0.01) |  |  | (-0.01 - -0.00) |
| 2nd SoE | -16.03** | -16.65** | -3.46** | -7.77** | -3.10* | 5.84** | 4.02** | 2.59** | -1.70** |
|  | (-17.36 - -14.69) | (-19.03 - -14.27) | (-4.71 - -2.20) | (-9.41 - -6.13) | (-5.70 - -0.50) | (4.77 - 6.91) | (3.19 - 4.86) | (1.01 - 4.16) | (-2.45 - -0.95) |
| 2nd SoE (days) |  | 0.02 | -1.05** |  | -0.13** | -0.69** |  | 0.04* | 0.37** |
|  |  | (-0.07 - 0.11) | (-1.18 - -0.91) |  | (-0.19 - -0.07) | (-0.77 - -0.61) |  | (0.00 - 0.08) | (0.34 - 0.40) |
| 2nd SoE^2 (days) |  |  | 0.01** |  |  | 0.01** |  |  | -0.00** |
|  |  |  | (0.01 - 0.02) |  |  | (0.01 - 0.01) |  |  | (-0.00 - -0.00) |
| 3rd SoE | -2.65** | -4.23** | -5.33** | 2.19* | -0.10 | -0.79 | 0.30 | 0.69 | 1.05* |
|  | (-4.49 - -0.82) | (-6.90 - -1.56) | (-7.95 - -2.71) | (0.55 - 3.84) | (-3.15 - 2.95) | (-3.87 - 2.28) | (-0.42 - 1.02) | (-0.24 - 1.62) | (0.12 - 1.97) |
| 3rd SoE (days) |  | 0.08* | 0.12 |  | 0.08* | 0.16 |  | -0.02 | -0.02 |
|  |  | (0.00 - 0.15) | (-0.02 - 0.25) |  | (0.01 - 0.16) | (-0.01 - 0.33) |  | (-0.04 - 0.01) | (-0.08 - 0.03) |
| 3rd SoE^2 (days) |  |  | -0.00 |  |  | -0.00* |  |  | 0.00 |
|  |  |  | (-0.00 - 0.00) |  |  | (-0.00 - -0.00) |  |  | (-0.00 - 0.00) |
| 4th SoE | -5.98** | -6.08** | -10.06** | -0.51 | 0.18 | -3.52** | 1.92** | 1.65** | 2.44** |
|  | (-8.78 - -3.18) | (-9.80 - -2.36) | (-12.97 - -7.14) | (-1.65 - 0.63) | (-0.61 - 0.97) | (-4.33 - -2.71) | (0.98 - 2.86) | (0.76 - 2.53) | (1.91 - 2.96) |
| 4th SoE (days) |  | -0.01 | -0.11 |  | -0.04* | -0.14** |  | 0.02 | 0.05* |
|  |  | (-0.08 - 0.07) | (-0.24 - 0.03) |  | (-0.07 - -0.01) | (-0.24 - -0.04) |  | (-0.01 - 0.04) | (0.01 - 0.09) |
| 4th SoE^2 (days) |  |  | 0.00 |  |  | 0.00 |  |  | -0.00 |
|  |  |  | (-0.00 - 0.01) |  |  | (-0.00 - 0.00) |  |  | (-0.00 - 0.00) |
| Observations | 13,455 | 13,455 | 13,455 | 13,455 | 13,455 | 13,455 | 13,455 | 13,455 | 13,455 |
| R-squared | 0.75 | 0.75 | 0.73 | 0.81 | 0.81 | 0.81 | 0.82 | 0.82 | 0.80 |

Note: The table presents coefficients with confidence intervals estimated from robust standard errors in parentheses. \*\* $p < 0.01$ , \* $p < 0.05$ . All models included the per-day confirmed infections in the past seven days, prefecture-level fixed-effects, prefecture-by-holiday fixed-effects, prefecture-by-weekdays fixed-effects, prefecture-by-month fixed-effects, and prefecture-by-year fixed-effects, and were weighted by population size. In addition, days since the start of the observation is included in Model (2), and additionally its squared term in Model (3). Full results are available upon request. A prefecture was classified as 'High' if the cumulative number of confirmed cases in that area was above the median of all prefectures within the study period. SoE = State of Emergency

**Appendix Table 15.** Trust in the government and mobility

|  | Retail and recreation | Supermarkets and pharmacies | Parks | Public transport | Workplaces | Residential area |
| --- | --- | --- | --- | --- | --- | --- |
| SoE | 76.45<br>(-41.48 - 194.38) | -27.30<br>(-110.54 - 55.93) | -82.41<br>(-298.51 - 133.68) | 94.84<br>(-34.03 - 223.72) | 53.22<br>(-79.82 - 186.26) | -21.84<br>(-69.12 - 25.43) |
| Trust in the government (Last week) | 0.41**<br>(0.28 - 0.55) | -0.00<br>(-0.10 - 0.09) | 0.49**<br>(0.24 - 0.73) | 0.65**<br>(0.50 - 0.79) | 0.21**<br>(0.06 - 0.36) | -0.18**<br>(-0.24 - -0.13) |
| SoE* Trust in the government | -2.48<br>(-5.56 - 0.60) | 0.60<br>(-1.57 - 2.77) | 1.88<br>(-3.76 - 7.52) | -2.96<br>(-6.33 - 0.40) | -1.65<br>(-5.12 - 1.82) | 0.73<br>(-0.51 - 1.96) |
| Covid cases (Last week: 100) | 0.96<br>(-0.85 - 2.78) | 3.90**<br>(2.62 - 5.18) | 2.95<br>(-0.37 - 6.27) | 2.70**<br>(0.72 - 4.68) | 2.58*<br>(0.53 - 4.62) | -0.18<br>(-0.91 - 0.55) |
| Observations | 577 | 577 | 577 | 577 | 577 | 577 |
| R-squared | 0.38 | 0.12 | 0.08 | 0.39 | 0.12 | 0.32 |

Note: The table presents coefficients with confidence intervals estimated from robust standard errors in parentheses. \*\* p<0.01, \* p<0.05. Trust in the government was obtained each month, and linearly interpolated to obtain the daily basis data. All models included holiday fixed-effects, weekdays fixed-effects, month fixed-effects, and year fixed-effects. SoE = State of Emergency

**Appendix Table 16.** Vaccination rates and mobility

|  | Retail and recreation | Supermarkets and pharmacies | Parks | Public transport | Workplaces | Residential area |
| --- | --- | --- | --- | --- | --- | --- |
| SoE | -7.22**<br>(-8.08 - -6.36) | 0.72**<br>(0.30 - 1.14) | -9.69**<br>(-11.66 - -7.71) | -6.35**<br>(-8.41 - -4.29) | -0.68<br>(-4.19 - 2.82) | 1.57**<br>(0.61 - 2.53) |
| Vaccination rate | 7.78**<br>(3.17 - 12.39) | 23.67**<br>(18.90 - 28.44) | -34.93**<br>(-58.29 - -11.57) | -5.84<br>(-16.98 - 5.30) | -10.74**<br>(-16.97 - -4.50) | -2.00#<br>(-4.11 - 0.12) |
| SoE*Vaccination rate | 22.39**<br>(16.29 - 28.48) | -4.83*<br>(-8.66 - -0.99) | 47.41**<br>(32.74 - 62.08) | 7.30<br>(-4.08 - 18.69) | -3.52<br>(-21.89 - 14.86) | -1.59<br>(-7.30 - 4.12) |
| Covid cases (Last week: 100) | -0.36**<br>(-0.48 - -0.25) | -0.12**<br>(-0.15 - -0.09) | -0.36**<br>(-0.47 - -0.24) | -0.31**<br>(-0.49 - -0.13) | -0.02<br>(-0.10 - 0.05) | 0.11**<br>(0.07 - 0.14) |
| Observations | 7,653 | 7,653 | 7,653 | 7,653 | 7,653 | 7,653 |
| R-squared | 0.81 | 0.59 | 0.69 | 0.77 | 0.80 | 0.83 |

Note: The table presents coefficients with confidence intervals estimated from robust standard errors in parentheses. \*\* p<0.01, \* p<0.05. Vaccination rates, which denote the number of those vaccinated at least once per population, were available between 12 April 2021 and 21 September 2021 on a daily basis for each prefecture. All models included holiday fixed-effects, weekdays fixed-effects, month fixed-effects, and year fixed-effects. Constants are not presented. SoE = State of Emergency

